# NIAGADS: A Comprehensive National Data Repository for Alzheimer’s Disease and Related Dementia Genetics and Genomics Research

**DOI:** 10.1101/2024.10.07.24315029

**Authors:** Amanda Kuzma, Otto Valladares, Emily Greenfest-Allen, Heather Nicaretta, Maureen Kirsch, Youli Ren, Zivadin Katanic, Heather White, Andrew Wilk, Lauren Bass, Jascha Brettschneider, Luke Carter, Jeffrey Cifello, Wei-Hsuan Chuang, Kaylyn Clark, Prabhakaran Gangadharan, Jacob Haut, Pei-Chuan Ho, Wenhwai Horng, Taha Iqbal, Yumi Jin, Peter Keskinen, Alexis Lerro Rose, Michelle K Moon, Joseph Manuel, Liming Qu, Flawless Robbins, Naveensri Saravanan, Jin Sha, Sam Tate, Yi Zhao, Alzheimer’s Disease Sequencing Project, Laura Cantwell, Jake Gardner, Shin-Yi Chou, Jung-Ying Tzeng, William Bush, Adam Naj, Pavel Kuksa, Wan-Ping Lee, Yuk Yee Leung, Gerard Schellenberg, Li-San Wang

## Abstract

NIAGADS is the National Institute on Aging (NIA) designated national data repository for human genetics research on Alzheimer’s Disease and related dementia (ADRD). NIAGADS maintains a high-quality data collection for ADRD genetic/genomic research and supports genetics data production and analysis. NIAGADS hosts whole genome and exome sequence data from the Alzheimer’s Disease Sequencing Project (ADSP) and other genotype/phenotype data, encompassing 209,000 samples. NIAGADS shares these data with hundreds of research groups around the world via the Data Sharing Service, a FISMA moderate compliant cloud-based platform that fully supports the NIH Genome Data Sharing Policy. NIAGADS Open Access consists of multiple knowledge bases with genome-wide association summary statistics and rich annotations on the biological significance of genetic variants and genes across the human genome. NIAGADS stands as a keystone in promoting collaborations to advance the understanding and treatment of Alzheimer’s disease.

## Introduction

Human genetics signals and functional genomic data holds great promise for nominating candidate therapeutic targets in complex diseases. Much progress has been made in the genetics of Alzheimer’s Disease (AD) since genome-wide association studies (GWAS) were introduced in the mid-2000s. Large collaborations such as the Alzheimer’s Disease Genetics Consortium (ADGC)^1^, International Alzheimer’s Genomics Project (IGAP) and population surveys such as UK Biobank^2^ increased sample size and statistical power. These studies^3–11^ have reported multiple high confidence, reproducible findings, confirmed the causal role of immunity, and helped clarify genetic (dis)similarities with other AD related neurodegenerative disorders. We see the following trends and challenges in advancing Alzheimer’s disease genetics: (1) Increasing sample size, especially for underrepresented populations, for sufficient statistical power in gene finding; (2) transitioning to rich phenotypes, biomarkers, and functional omics beyond case/control status; (3) developing novel statistical methods, analysis platforms and tools for rare variant discovery; (4) generating new approaches to translate genetic findings to mechanisms and pathways. Developing accurate, population-specific disease risk models will allow identification of high-risk individuals for trial recruitment and development of sensitive early diagnostics. To this end, researchers need a large, high-quality AD genetics data resource with rich phenotypes and functional genomic data.

NIAGADS is the National Institute on Aging (NIA) designated national data repository for AD and AD-related dementia (ADRD) human genetics research. Supported by a U24 cooperative agreement with the University of Pennsylvania since 2012, NIAGADS functions as a one-stop access portal for AD genetics, managing and sharing genetic data and findings with the research community. The funding coincided with the announcement of the Alzheimer’s Disease Sequencing Project (ADSP), an NIA strategic initiative to analyze complete genomes of AD patients and cognitively normal controls from diverse populations to find novel genetic variants modulating AD risk. NIAGADS is the data coordinating center for ADSP. See Figure 1 for the timeline of NIAGADS and ADSP data growth.

**Figure 1.**
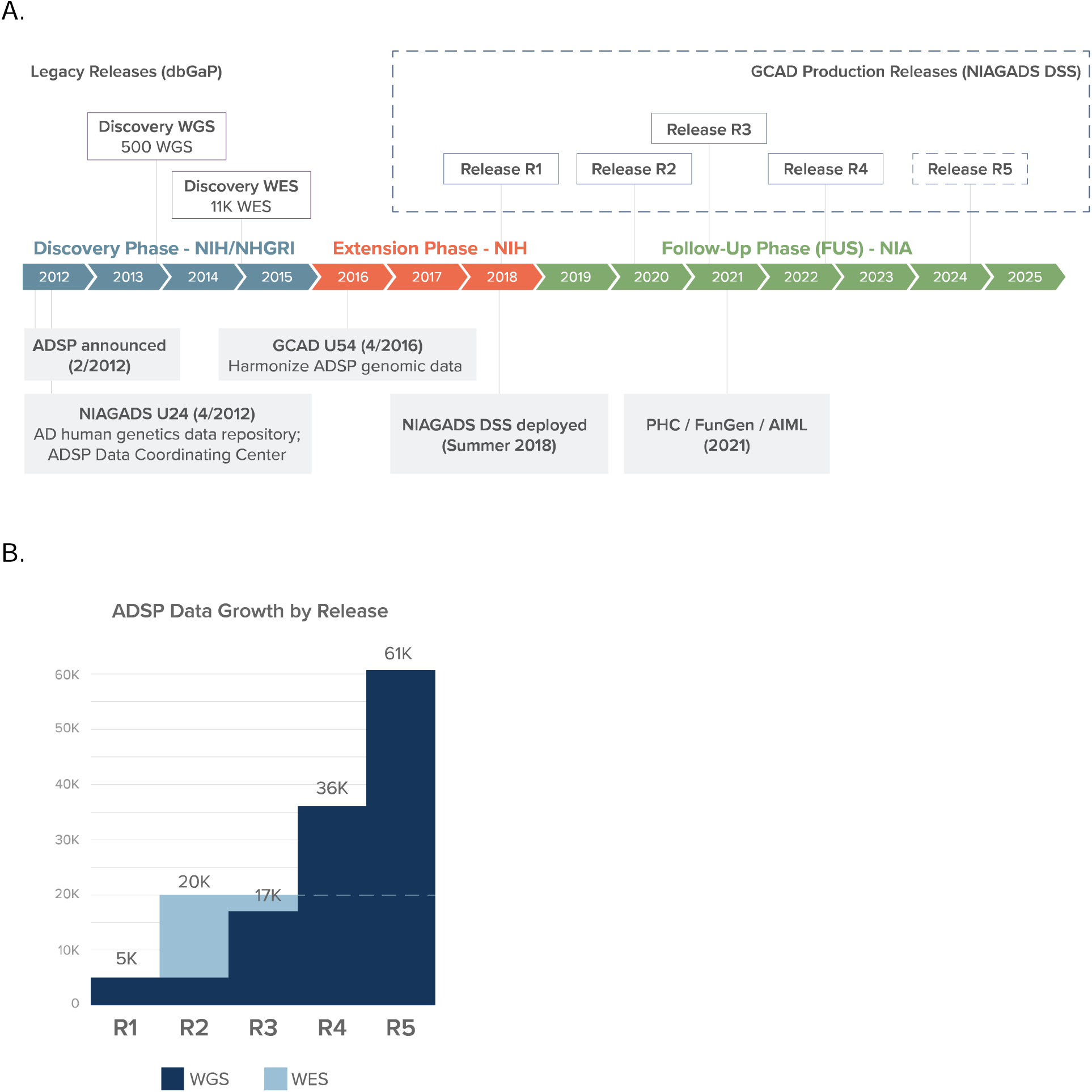
(a) NIAGADS and ADSP Timeline. (b) ADSP Data Growth by Releases.

NIAGADS has three major objectives: (1) Curate, update, and disseminate a high-quality data collection for genetic/genomic research of AD/ADRD; (2) Develop secure informatics infrastructure for data management, dissemination and use; (3) Support AD genetics data production and analysis activities. This article reports the available datasets, design principles, and IT infrastructure of NIAGADS as a valuable resource for the research of Alzheimer’s disease and human genetics in general.

## Results

### Overview

NIAGADS provides two web-based platforms for users to explore and access its rich and complex data collection (Figure 2). Individual datasets are released via the NIAGADS Data Sharing Service (DSS) website. Sensitive datasets such as individual GWAS genotype data and ADSP whole genome sequences require a review process mandated by NIH and are available by a Data Access Request (DAR) process that is compliant with NIH Genome Data Sharing (GDS) Policy. Datasets that are not sensitive can be downloaded directly via the DSS Open Access data portal. AD genetics knowledge and genomic annotation are available via NIAGADS Open Access, an integrated suite of genomics knowledge bases coupled with web interfaces for querying available data, viewing gene and variant reports, and allows much of the annotation data to be downloaded directly.

**Figure 2.**
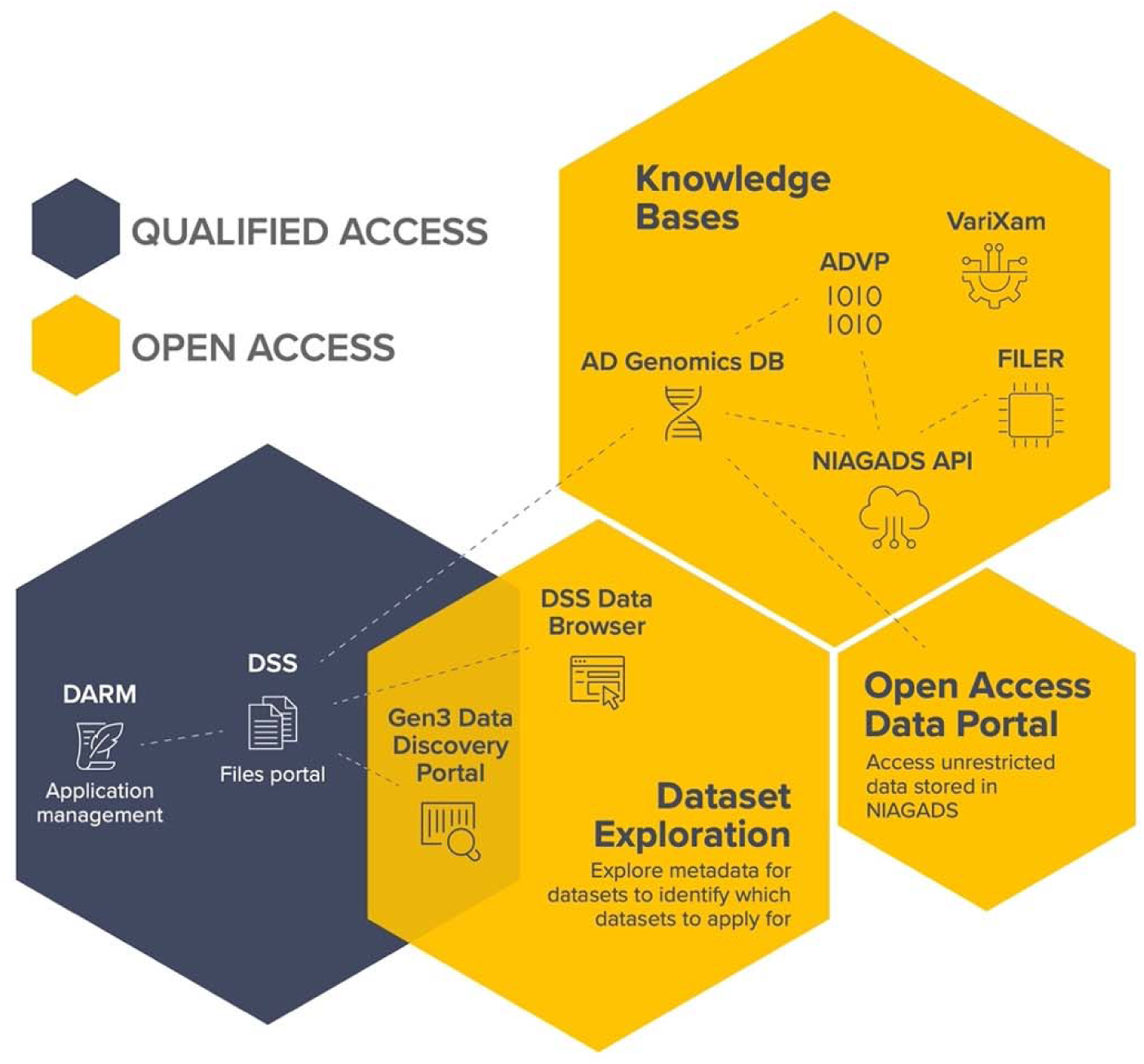
Layout of the NIAGADS platforms. NIAGADS shares controlled-access human genetics/genomics data and open-access genomic knowledge through an integrated system of interoperable data access platforms. The dual approaches facilitate broad data sharing while ensuring compliance with ethical and privacy standards.

### AD genetics datasets

As of June 2024, NIAGADS has 125 distinct datasets of various genomic data types for more than 209,000 samples (Figure 3). The primary data types hosted at NIAGADS are human genetic data from GWAS SNP arrays and imputations, and whole-exome/whole-genome sequencing data (WES/WGS). Both individual level data and summary statistics are available. Genetic data are connected with clinical phenotypes including Alzheimer’s disease case/control diagnosis, age at onset/last visit/death, sex and population (based on cohort reports). Other data types include functional omics, fluid biomarkers, aggregated imaging data (such as volumetric data for brain regions of interest), and summary statistics. For a significant (and increasing) subset of these subjects, fully harmonized phenotype data, developed by the ADSP Phenotype Harmonization Consortium (PHC), are available through NIAGADS. Datasets are governed by more than 200 institutional certifications that provide informed consent research use limitations determined by the Institutional Review Boards (IRBs) governing these studies. Figure 3b shows the location of cohorts that contribute to NIAGADS data collection. These datasets are used widely by the research community around the world. As of June 2024, NIAGADS supported more than 500 unique data requests from qualified investigators and is officially acknowledged in over 330 articles (Figure 3a).

**Figure 3.**
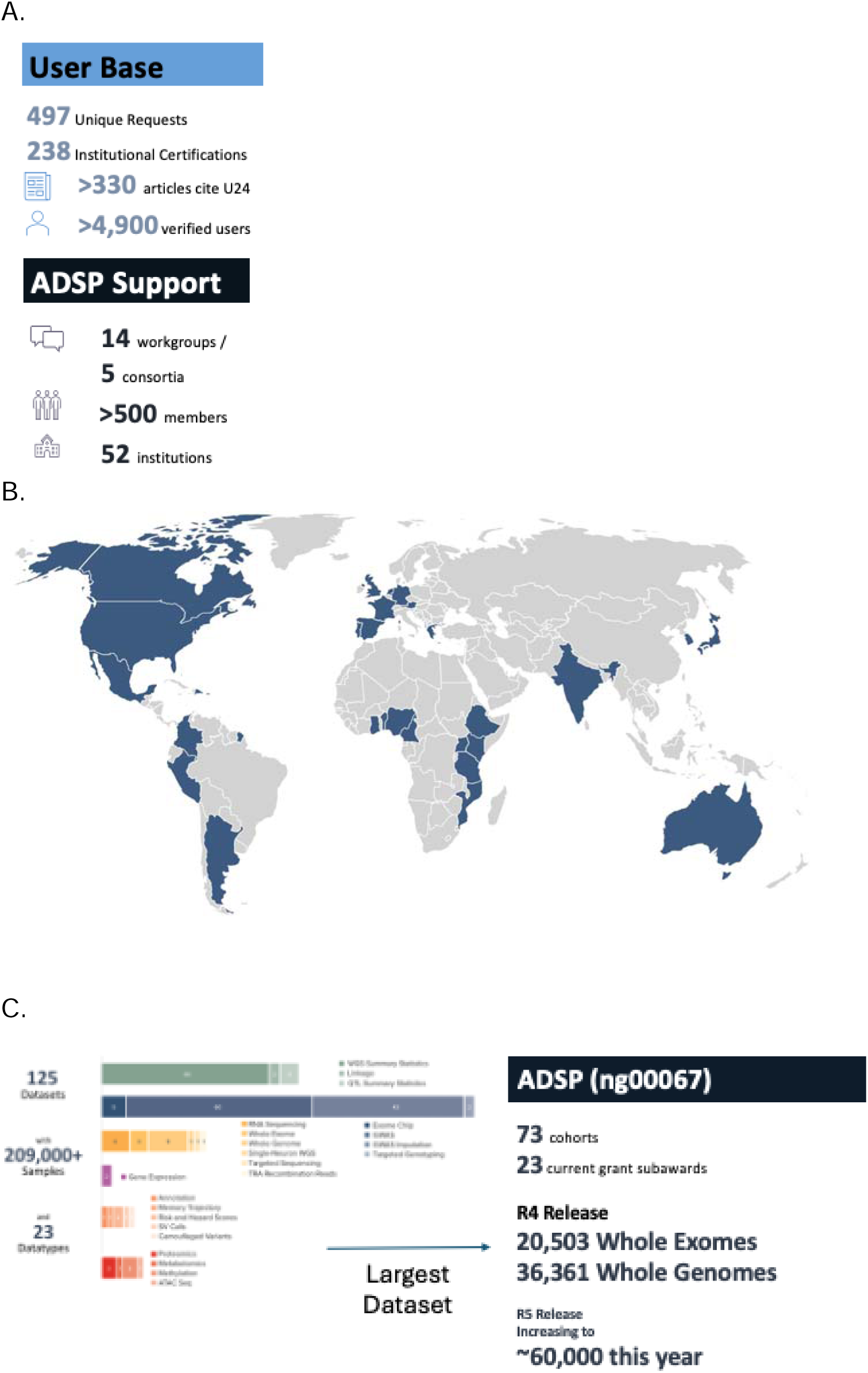
NIAGADS at a glance. (a) NIAGADS by the numbers. (b) A global map of cohorts contributing to the NIAGADS data collection. (b) The NIAGADS Data Collection breakdown by data types.

The largest data in NIAGADS in terms of data size are whole genome and whole exome sequence data as well as derived data (e.g. structural variant calls) and associated phenotypic data from the ADSP. These data are produced by participating cohorts, infrastructure grants, and workgroups. Currently NIAGADS works with 73 cohorts in ADSP and will add more cohorts into ADSP in future releases.

NIAGADS offers a comprehensive collection of genomic data types for Alzheimer’s Disease and Related Dementias (ADRD) research. The majority of data in NIAGADS are relevant to human genetics/genomics research, with datatypes such as summary statistics, GWAS SNP array data (with imputation using very large reference panels), exome chips, WGS/WES data, and associated clinical phenotype data. For some of the cohorts managed by NIAGADS, NIAGADS also hosts omics data such as RNA-Seq, proteomics, methylation, and metabolomics data.

See Figure 3c for a breakdown of data stored in NIAGADS as of July 2024.

### ADSP data

NIAGADS started sharing ADSP sequence data with the research community in 2018. ADSP continues to generate WGS data from cohorts selected by the National Institute on Aging. New genomic data are passed to the Genome Center for Alzheimer’s Disease (GCAD) to generate compressed read alignment files (in the CRAM format) and individual genotype calls (in the genomic VCF or gVCF format) which are then released in batches after sample level quality control is completed. In addition, ADSP releases fully harmonized genetic data as cumulative joint genotype calls (project-level VCF files) by GCAD for every observed variant across all previously released and new genomes. Between these major releases, ADSP also releases additional information including in-depth quality control results, population structure principal component estimates, variant annotations, and structural variant calls. As of June 2024, ADSP has released four major freezes (R1: 4,789 genomes; R2: 20,503 exomes; R3: additional 12,116 genomes and joint genotype calls of 16,905 genomes; R4: additional 19,456 genomes and joint genotype calls of 36,361 genomes – see Figures 1b and 4) across 13 versions. All genome sequences have been processed using a GATK-based processing pipeline^12^ against the human genome reference 38 genome build. All exome sequences have been processed using the same pipeline that accounts for differences in capture region designs from 10 different exome capture kits ^13^ to retain as many variants as possible. R2 WES data includes 8.2 million autosomal variants, and R4 WGS data includes more than 438 million variants. Starting with R4, ADSP has more genomes from underrepresented non-European groups than genomes from European participants. ADSP is currently working on the next R5 release (fall 2024) which will bring the total number of genomes to more than 60,000 from 57 different cohorts.

**Figure 4.**
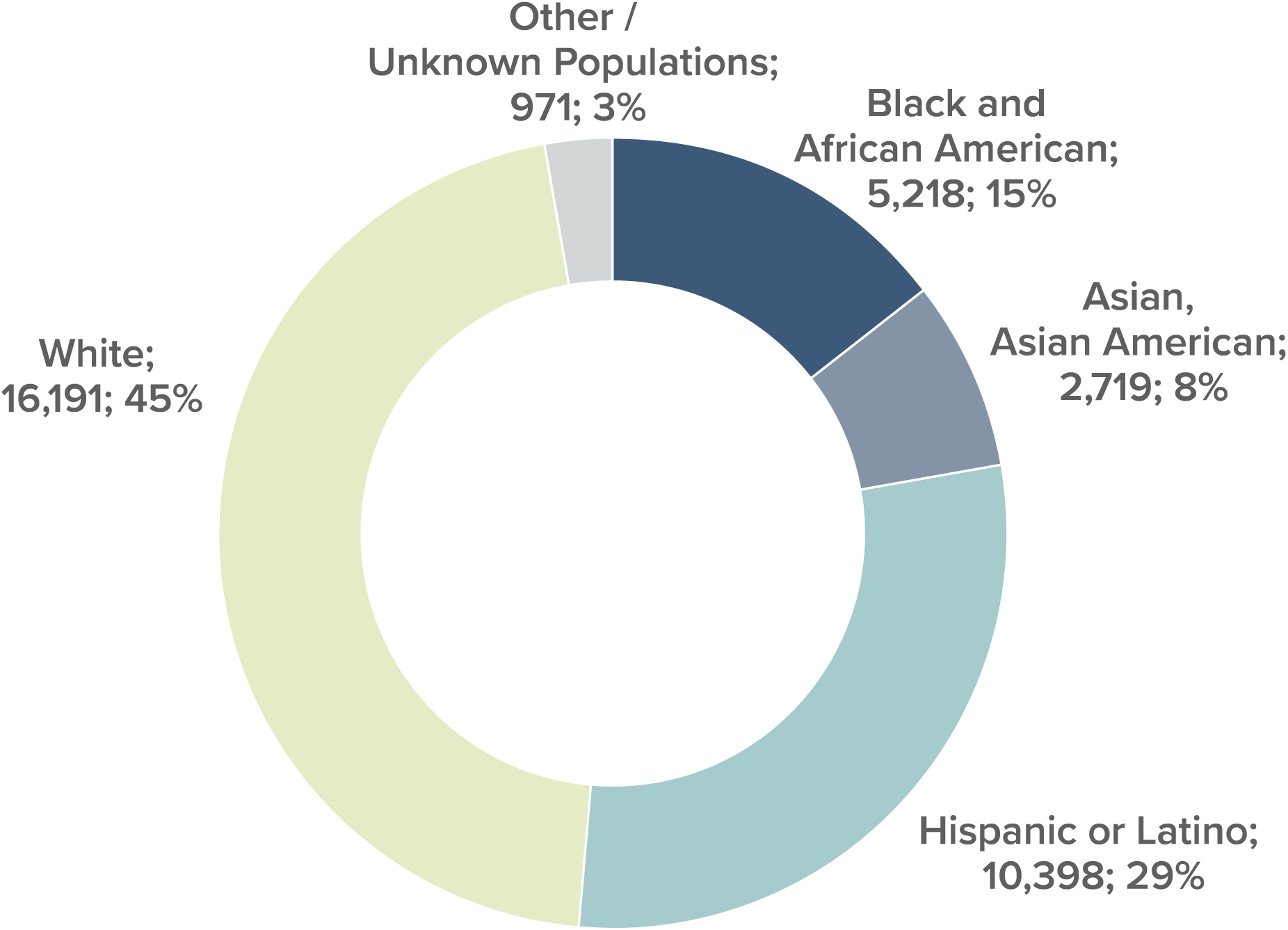
ADSP Release 4 whole genome sequence data by cohort-reported population.

### ADGC GWAS data

NIAGADS releases GWAS SNP array data submitted by the ADGC. ADGC assembles AD cohorts, collects GWAS SNP array data, and performs data harmonization, quality control, and imputation using the latest reference panels (TOPMed-r2 panel consisting of 97,256 genomes and 308 million variants across diverse populations).

ADGC also genotypes DNA from subjects recruited by more than 30 NIA Alzheimer’s Disease Research Centers (ADRCs)^14^ as part of the National Alzheimer’s Coordinating Center (NACC)^15^ data collection. In Spring 2024, NIAGADS released 15 batches of the NACC sample genotyping array datasets generated by ADGC (Table 1). The dataset consists of 29,694 subjects with TOPMed-r2 imputed genotype data. All data have been mapped to the human reference genome 38 and sample ID mappings to ADSP genomes are included so researchers can link across the two datasets.

**Table 1.**
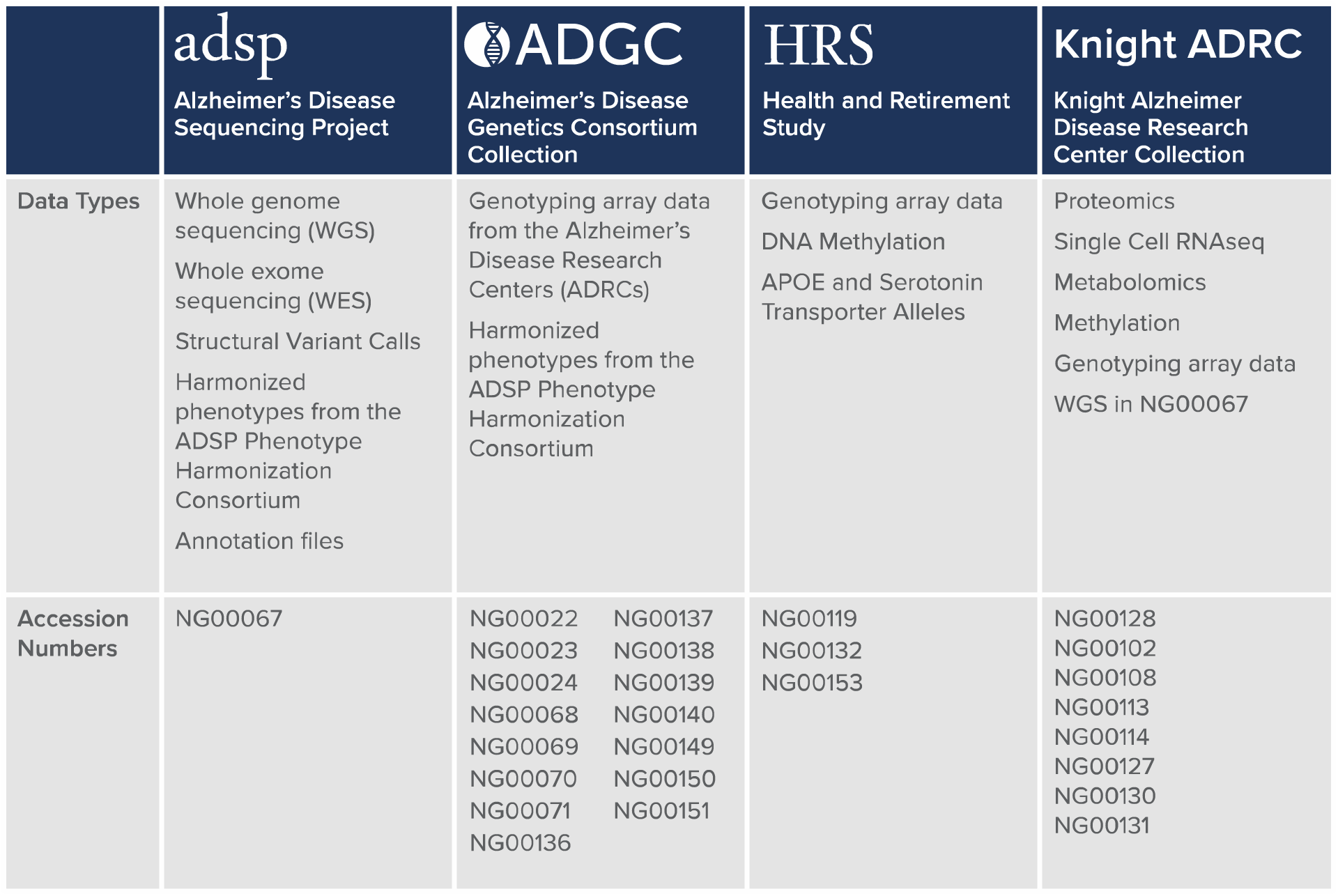
Featured datasets: ADSP, ADGC, HRS genomics data, and Knight ADRC Genetics Collection.

### Other human genetics datasets

NIAGADS houses many other datasets such as Health and Retirement Study (HRS) GWAS and Knight ADRC collection datasets (Table 1), and data derived from NIAGADS datasets such as genome-wide summary statistics submitted by investigators from published studies. Users can access datasets with special research use limitations (e.g. sensitive population or required by the original informed consent) by submitting data access requests. NIAGADS also has data for participants with other types of dementias like Parkinson’s disease (PD), frontotemporal dementia (FTD), Lewy body dementia (LBD), and progressive supranuclear palsy (PSP) (Figure 2). Of note, NIAGADS hosts whole genome sequences of 1,992 samples with PSP as part of the ADSP release.

### NIAGADS Data Sharing Service (DSS)

Most of the datasets housed within NIAGADS require users to submit a data access request (DAR) as there are special research use limitations as required by the original informed consent. Whenever possible, NIAGADS makes summary statistics without such limitations available for direct download or through the NIAGADS Open Access resources. NIAGADS DSS is a qualified access data sharing platform following relevant NIH and federal policies. Much of the platform design and organization is based on the dbGaP website to ensure policy compliance and familiarity to the research community. NIAGADS DSS is compliant with FISMA Moderate risk level requirements^16^ built on the Amazon Web Services^17^ (AWS) cloud platform. DSS relies on the NIH eRA grant management system to validate user identities.

NIAGADS supports and implements the NIH Genomic Data Sharing (GDS) policy (July 2024 update)^18^ and is listed as a NIH security best practices repository (https://sharing.nih.gov/accessing-data/NIH-security-best-practices). All cohorts submit a completed Institutional Certification approved by the cohort’s institutional Review Board (IRB). The IRB reviews the original consent forms and determines the research use limitations using the GDS framework, including permission to use the data for research outside AD/ADRD and permissibility for use by for-profit organizations. If the cohort has multiple versions of informed consent, the cohort needs to specify which research use limitation applies to each participant in the consent form. Per GDS policy, certain cohorts may specify research use limitations for genomic summary results (e.g. populations of special status such as tribal nations in the United States) and will require the requestor submit a formal data request.

The cloud-based DDS platform consists of three components. The *File Repository* includes a web-based data portal front end that allows users to browse and filter all files in each dataset and links to the file objects on the Simple Storage Service (S3) buckets on the Amazon Web Service. The *DSS website* contains detailed information about submitting and managing Data Access Requests (DARs), a catalog of all datasets and detailed metadata such as cohorts, acknowledgements and grant numbers, Digital Object Identifiers (DOIs), sample size, file manifests (with MD5 checksum) and version/release history. The DSS website lists all approved DARs with succinct research use summaries and has a tool that generates the acknowledgement statement from a list of dataset accession numbers.

The *Data Access Request Management (DARM)* system is the core of the DSS platform that manages user and file access permissions. Users log into DARM using their NIH eRA login credential to submit new DARs and manage existing requests. The DAR submission interface is similar to the dbGaP system with additional features developed based on user feedback such as a DAR administrator role, which allows the requesting investigator to designate a preparer to enter request information before submission by the investigator. The DARM system also implements many features to support the Data Access Committee. The NIA ADSP Data Access Committee (NADAC) consist of NIH program officers who are federal employees. The DARM system allows NADAC members to review all requested materials, vote to approve for each requested dataset by each research use limitation level based on the research use statement and requesting institution. Review outcomes include approving a DAR, requesting additional information or correction, or rejecting the DAR. All approved DARs are required to submit annual renewal requests with progress reports (e.g. summary of findings and publications) to retain access. See Figure 5 for the DAR review process.

**Figure 5.**
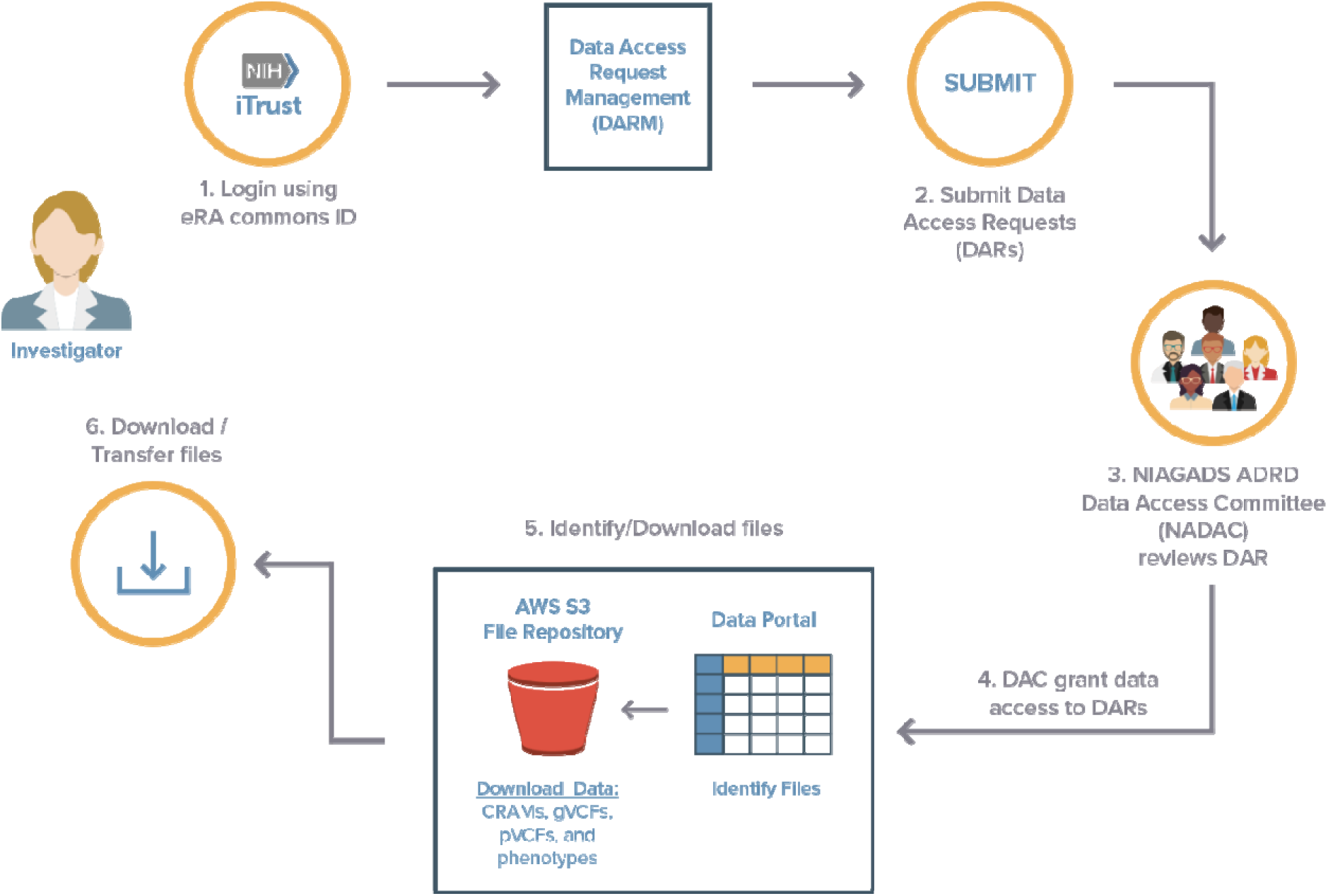
Overview of the Data Access Request (DAR) submission and review process in the NIAGADS Data Sharing Service (DSS). Users start by browsing the meta data for available datasets and access documentation for submitting or applying for datasets. 1. Using an eRA commons ID, users log into the DSS platform where they can submit look at the applications or datasets they have available to them. 2. In the DARM, users can submit, revise, renew, or close data applications. 3. Approving officials can review user applications for applicability for each consent level of each dataset users applied for, which informs what files users can see in the files portal. 4. Users are notified their application is approved. 5. In the Files Portal, users can see files they have access to based on their approve application. 6. Users transfer or download the files from AWS.

### NIAGADS Open Access

NIAGADS hosts many non-sensitive analysis summaries and genome annotations that do not require a special request and review process. The NIAGADS Open Access (Figure 6) platform facilitates access to these datasets via direct file download from the Open Access data portal on the DSS website (or through AWS) and other resources and services: AD GenomicsDB^19^, ADVP^20^, FILER^21^ and VariXam.

**Figure 6.**
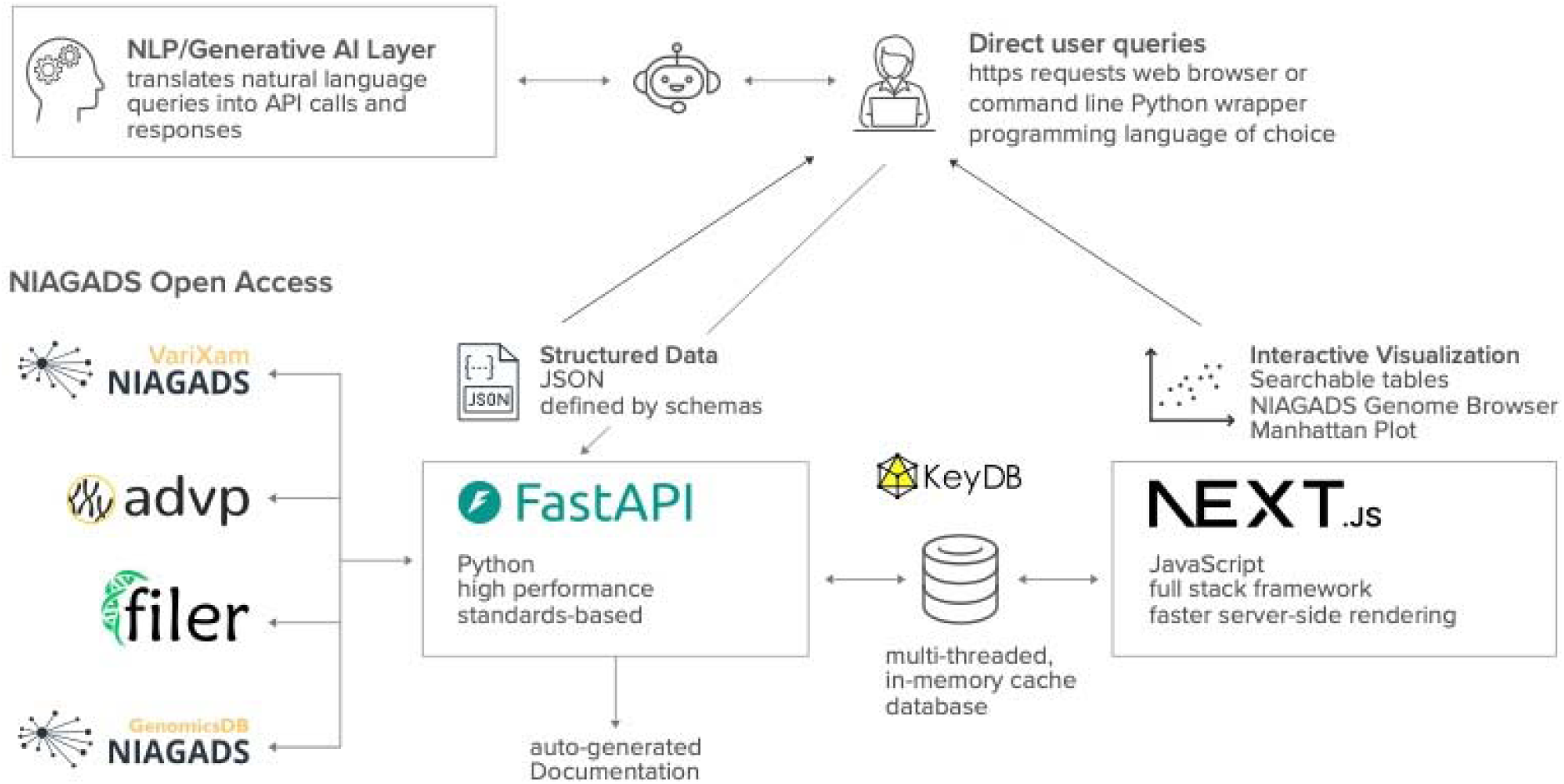
NIAGADS Open Access Knowledge Bases and API architecture. See Online Methods for technical details.

*The NIAGADS Alzheimer’s Genomics Database (AD GenomicsDB)* is a public knowledge base for the integration of AD genetic findings, genomic annotations, analysis summaries, and ADSP variant information. As the core of the NIAGADS Open Access framework, the AD GenomicsDB is powered by the PostgreSQL relational database engine, optimized for large-scale queries and scalable data integration. All records are assigned unique stable IDs for cross-version compatibility and harmonized for compatibility with ADSP data, ENSEMBL^22^ , dbSNP^23^, and other genomic databases.

The AD GenomicsDB hosts a wide range of information of biological significance associated with variants, genes, and genomic intervals. More than 438 million annotated single-nucleotide polymorphisms (SNPs) and insertions/deletions (indel) from the ADSP R4 data freeze are available with key information such as observed alleles, population frequency (from gnomAD^24^, 1000Genomes 30x^25^, and the NIH ALFA project^26^, GWAS findings (from GWAS Catalog^27^, ADVP, and more than 60 genome-wide GWAS summary statistics in NIAGADS), co-located genes, effect on the coding sequence or predicted functional impact, loss of function, and CADD deleteriousness scores (using the ADSP Annotation Pipeline). AD GenomicsDB assigns unique variant identifiers by chromosomal coordinates and allelic variants, ensuring accurate mapping of risk-association statistics. Gene models are linked to various identifies such as ENSEMBL IDs and NCBI Gene IDs and linked with key information from external databases including pathways (e.g. KEGG^28^ and Reactome^29^), functional annotations (e.g. Gene Ontology^30^), protein information (UniProt^31^) and clinical significance (OMIM^32^).

The AD GenomicsDB website is built using the VEuPathDB Web Development Kit^33^ to allow for a responsive interface and gene and variant report pages customized for AD human genetics research. Variant report pages provide context views of the genomic region such as linkage disequilibrium (LocusZoom^34^). Finally, researchers can generate genome browser views using the AD GenomicsDB genome browser. Powered by IGV.js^35^, users can view various annotations in the given genomic region, add their own tracks, and interact with the data records to gain insights into genetic associations and functional activities and their interactions.

*The AD Variant Portal (ADVP)* provides a curated collection of statistically significant genetic associations for AD and related dementia in the research literature. The first version of ADVP, released in 2021, includes almost 7,000 genome-wide significant genetic associations from 125 ADGC publications. These signals come from 1,800 distinct variants in more than 900 loci and are associated with disease risk, expression quantitative traits, age at onset, biomarkers, neuropathology, and other relevant outcomes. All records are harmonized for variant rsIDs, allele coding, and genome coordinates (human reference genome builds hg37 and hg38) to ensure compatibility with ADSP data and analysis results. ADVP also captures relevant metadata including study population (cohort-reported), sample size, imputation panel, and publication. Users can search these records by publication title, gene name, and variant IDs, visualize the distributions of records on the human genome ideogram using the ADVP variant viewer, or follow URL links to access additional information in the AD GenomicsDB or PubMed database. The next version of ADVP will add 10,000 more association records from >450 new articles.

*The Functional genomics repository (FILER)* was designed to address the challenge of making large queries across different public sources of functional genomic annotations stored as “tracks” per genome browser jargon. Each track consists of between thousands and millions of genomic intervals with functional significance, including tissue-specific regulatory elements, transcription factor binding activity, chromatin states, genetic regulation (eQTL, sQTL), and chromatin conformation. Currently FILER has more than 70,000 tracks and more than 17 billion records for both hg37 and hg38 builds. The data come from more than 20 data sources such as ENCODE^36^, GTEx^37^, FANTOM5^38^, and NIH Roadmap Epigenomics^39^, and cover over 1,100 distinct tissue/cell types. FILER provides metadata such as tissue or cell type and experimental assay that are harmonized using ontology standards. Researchers can freely download all tracks in FILER or access individual records via its website. FILER provides a scalable query interface that can cross-compare all tracks and records against a user-provided list of genomic intervals and return all overlaps. This allows users to quickly identify functional significance and tissue context for genomic regions of interest, and generate large data feature matrices to be used in association tests, machine learning and artificial intelligence.

*VariXam* is a quick access viewer for assessing the quality of genetic variant calls in ADSP whole exome and whole genome data. Developed using a React framework for the frontend and PostgreSQL for the backend, VariXam facilitates efficient querying by gene names, coordinates, genomic regions, or variant rsIDs. The database encompasses all four major ADSP freezes, ensuring comprehensive data coverage for genetic variant analysis.

### FAIR compliance and community engagement

Introduced in 2016, the FAIR principles (Findable, Accessible, Interoperable, Reproducible)^40^ have become the ubiquitous guidelines for data repositories and knowledge bases. NIAGADS follows the FAIR principles when designing websites and databases, data sharing infrastructure, data curation workflows, and procedures for engaging and supporting researchers (Table 2). NIAGADS implements common standards in genetics and genomics, widely accepted best practices and software tools for processing genomic data, and community support. Beginning in 2023, NIAGADS now assigns Digital Object Identifiers^41^ (DOI) to each dataset, facilitated by NIH’s membership in the international DataCite consortium^42^. DOIs are persistent identifiers and will enhance FAIR compliance.

**Table 2.**
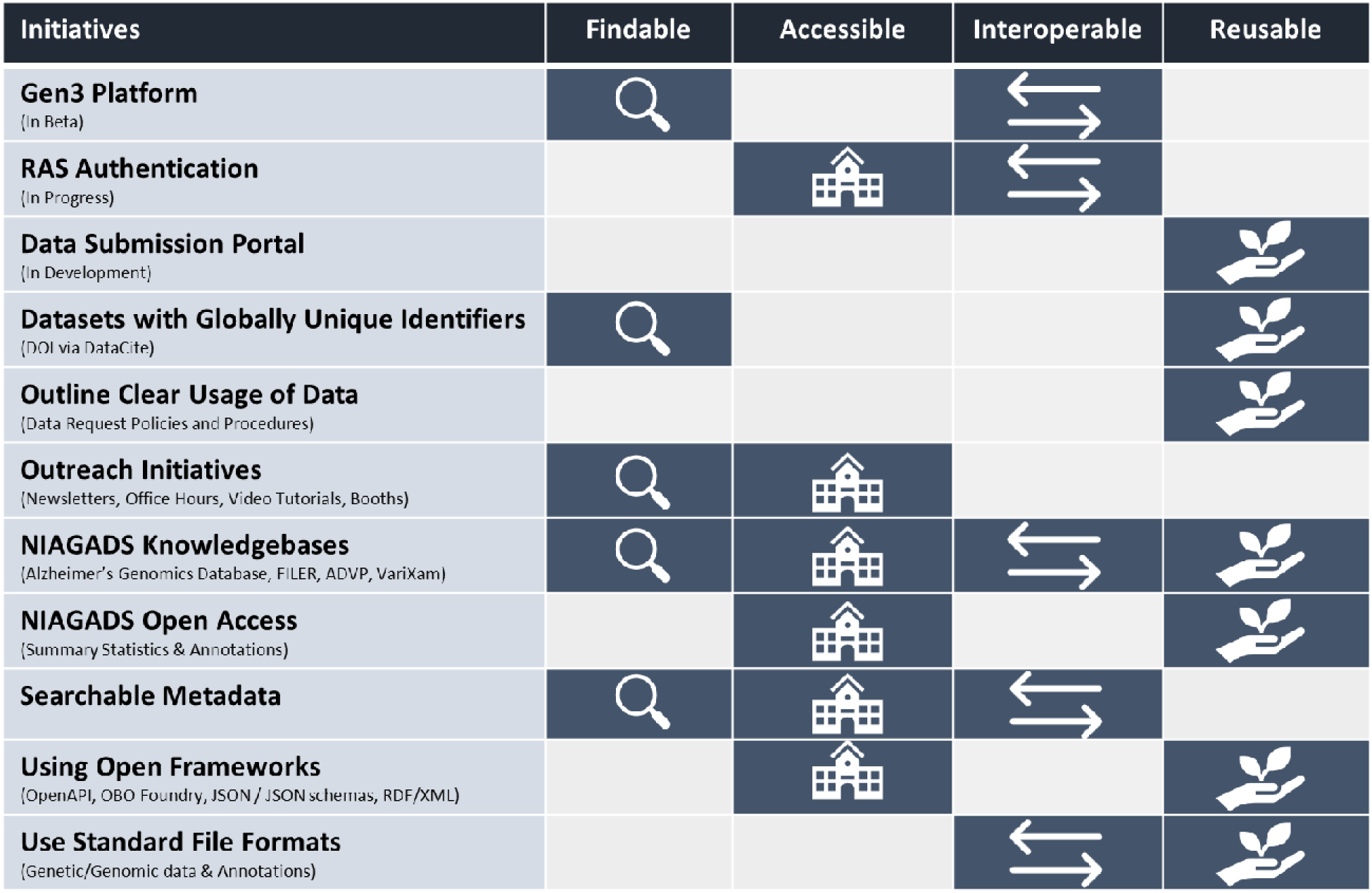
NIAGADS features in support of the FAIR principle (Findable, Accessible, Interoperable, Reproducible).

To complement these FAIR features, NIAGADS has an active community outreach and engagement program. In 2023 NIAGADS held 3 online office hours and developed video tutorials for GenomicsDB, and hosted booths at major conferences such as Alzheimer’s Association International Conference (AAIC), American Society for Human Genetics (ASHG), and Society for Neuroscience (SfN). NIAGADS has a user support web page with detailed documentation and frequently asked questions (FAQs), and a ticketing system for user requests. NIAGADS maintains news posts on the ADSP website with latest findings, data releases, and investigator profiles. NIAGADS has been releasing 3 joint NCRAD/NIAGADS newsletters each year. In May 2024, NIAGADS started releasing monthly newsletters. Table 2 summarizes how NIAGADS implements FAIR compliance.

### Data production, curation and partnership

Modern GWAS designs (SNP array or whole genome sequencing) often require many cohorts to obtain very large sample sizes to reach better statistical power. Thus, production and assembling of data for these projects require substantial planning and coordination beyond simply receiving data from contributing cohorts. As the data coordinating center for ADSP, NIAGADS achieves these goals through meticulous planning and close interactions with major contributors, producers, and stakeholders. NIAGADS implements a single data flow path (Figure 7) for receiving, processing and sharing data. All data coming into ADSP are sent to NIAGADS so data transfer agreements can be set up to ensure all subsequent data flows and sharing are compliant with the original informed consent. NIAGADS executes ADSP data production by routing raw and intermediate data to designated data processors with domain expertise and capacities and receiving processed data products. The single data flow path design ensures consistency, legitimacy, transparency, and reduces human errors.

**Figure 7.**
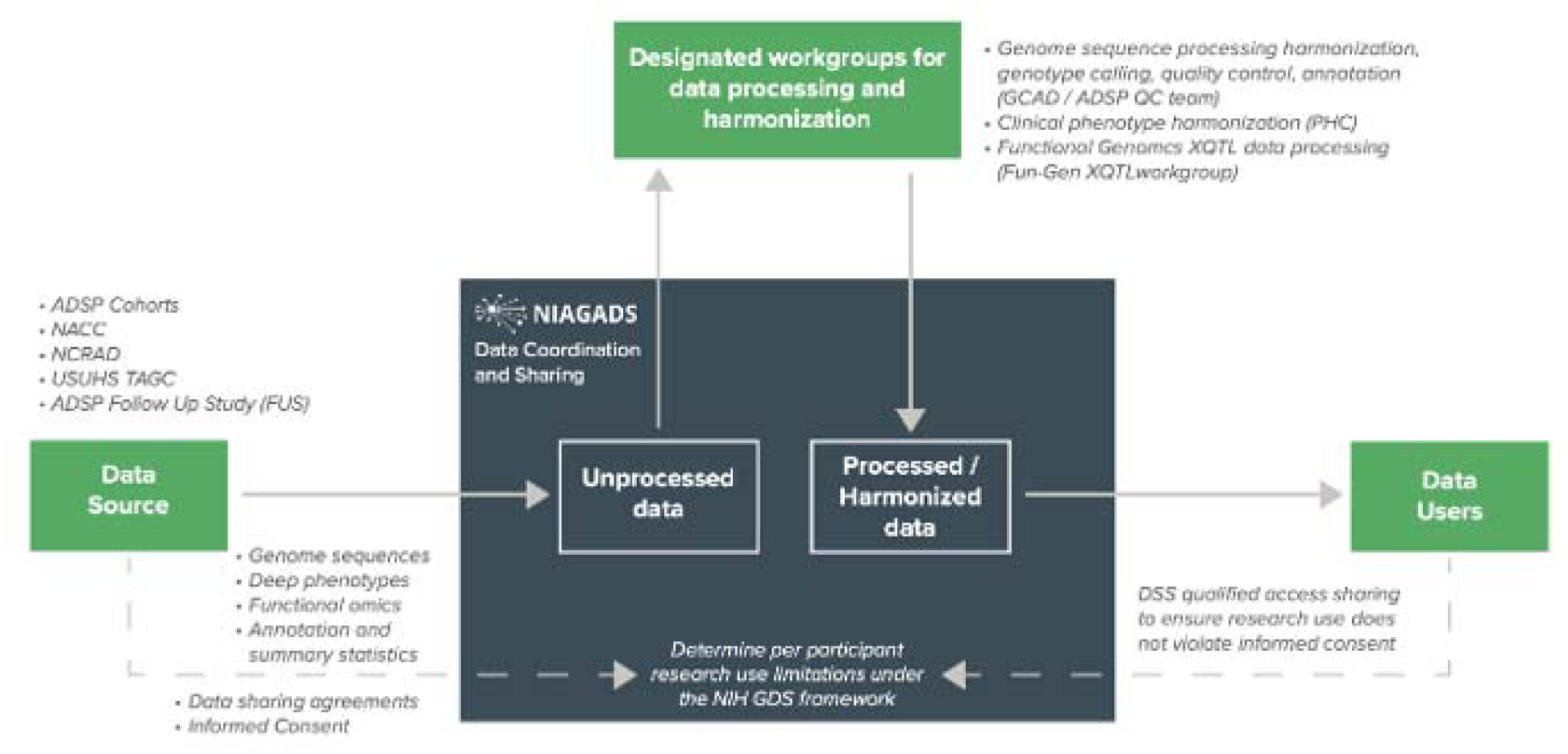
Flow diagram of the ADSP data production process. A unified data flow allows NIAGADS to manage and move files during the process and track the progress of all data processing and sharing activities, while ensuring compliance with NIH Genomic Data Sharing (GDS) policy. A straightforward and centralized coordination process is easy to understand and facilitates communication with all contributors. Data are dispatched to designated domain experts by data types.

The ADSP data production process begins when a new cohort joins the program. NIAGADS registers the cohort, collects research use limitations using the GDS Institution Certification form, and sets up data transfer agreements that authorizes NIAGADS to direct data to production partners and eventually share with qualified investigators through DSS. Currently NIAGADS works with 73 ADSP cohorts (and growing) from 31 countries across 6 continents governed by 21 different levels of research use limitations. These cohorts either provide completed whole genome sequence data in the form of raw sequencing reads or submit DNA to the National Centralized Repository for Alzheimer’s Disease and Related Dementias^43^ (NCRAD), an NIA-designated national biorepository operated by Indiana University. NCRAD performs quality control and plates the DNA for sequencing by one of the ADSP sequencing partners such as The American Genome Center (TAGC) at the Uniformed Services University of the Health Sciences (USUHS) or the John P. Hussman Institute for Human Genomics (HIHG) at the University of Miami School of Medicine. Raw sequencing data are returned to NIAGADS and then routed to the Genome Center for Alzheimer’s Disease (GCAD) for read mapping, variant calling, quality control, and documentation. Cohorts also submit phenotypes to NIAGADS, which relays to the Phenotype Harmonization Consortium^44^ for phenotype harmonization. NIAGADS works with NIA program officers, GCAD, sequencing partners, the ADSP Follow-Up Study coordinating team, and PHC to plan release dates and the final list of cohorts for each major freeze.

The same coordinating process is used to coordinate structural variant calling for all ADSP WGS data (processed by GCAD and the ADSP structural variant workgroup), and the multi-omics quantitative trait linkage (XQTL) project in the ADSP Functional Genomics Consortium (processed by GCAD and the XQTL workgroup). In addition, NIAGADS supports other components of the ADSP project, including the AI/ML consortium (common benchmark datasets for AI and machine learning algorithms), the gene verification committee (ADSP website hosts a list of AD susceptibility loci and genes with high confidence), and the ADSP Executive Committee/Cross-Consortium Communication Committee (ADSP website and project management support). NIAGADS also interoperates with other NIA infrastructure including the National Alzheimer’s Coordinating Center (NACC - returning sequencing and GWAS data to individual ADRCs), NCRAD (sample and genetic data availability across the two repositories), AD Knowledge Portal (the Agora website^45^ connects recommended therapeutic target genes to individual gene report pages in the AD GenomicsDB), and the AD NeuroImaging Initiative^46^ (ADNI; federated data sharing request process via the Laboratory of NeuroImaging (LONI) at the University of Southern California).

## Discussion

The design philosophy of NIAGADS is a natural consequence of the idiosyncrasies with studying Alzheimer’s Disease genetics. Although Alzheimer’s disease is a common ailment among seniors, recruiting large numbers of Alzheimer’s disease patients for research remains difficult. Even if neuroimaging is not used, proper diagnosis is still expensive, time consuming and requires special expertise in neurology or neuropsychiatry that is rarely available at community level primary care. AD remains stigmatic, especially among underrepresented minorities. Retaining AD patients in longitudinal studies are challenging and burdensome on the patients and caregivers but essential for the slowly converting and progressing disease. The only way to obtain hundreds of thousands of participants for human genetics research is through collaborations with all available cohorts, most of which would only comprise several hundred participants, some enrolled decades ago. Data collection procedures and diagnostic approaches have changed over years as our understanding of the disease evolved. Careful planning and coordination for phenotype and genotype harmonization and consistent adjudication is essential, time-consuming, unavoidable, but adds great value to the research community.

The DSS qualified data access process provides legal protection for researchers. A complete implementation of the NIH Genome Data Sharing policy properly captures research use limitations specified in the original informed consent for every participant and ensures that their data will only be used within the scope and by organizations that were agreed upon when they were recruited in the study. The whole process as defined in the GDS policy is complex and resource intensive, although continuous work with the data access committee to streamline the process is ongoing. Note that a single request allows investigators to access all the data in ADSP comprising decades of investment by the US government and the cumulative accomplishments by thousands of researchers. Proper protection is necessary to retain the trust granted by the participants when they signed on to these cohorts.

As mentioned earlier, DSS relies on the NIH eRA to verify data requesters and their institutions. As research institutions around the world register their investigators and signing officials with the NIH eRA platform, NIAGADS can verify if the institution is eligible to access qualified access NIH data (e.g. dbGaP), if the investigator is currently employed by the institution and in good standing, and if the signing official has legal authority to represent the institution. An important legal concept is that all data transfers are between institutions and not with the investigators directly, just like research grants are issued by the federal government to the investigator’s institution, not to the investigator. This is required so data protection requirements can be legally enforced. The validation process is especially important when data requests come from commercial entities and/or institutions outside the United States.

Coordination and direct support by NIAGADS remain critical to the ADSP production process since it started in 2012. The richness and complexity of ADSP data means every analysis will have its own quality control challenges and study design considerations. Nonetheless, most researchers will want genotype calls and not the massive raw data or detailed quality information. For this reason, NIAGADS releases compact versions of joint calls, with detailed quality tags removed to reduce file size, and preview versions of genotype calls after sample quality checks are completed. ADSP is composed of harmonized genetic and phenotypic data from studies with very different designs and missions, making it is impossible to define a single definitive version that works for all analysis objectives. To this end, it is essential that the data come with proper documentation and critical metadata. Additionally, NIAGADS has developed quick guides and scripts for phenotype integration for users to better understand the rich dataset.

Currently in beta, the NIAGADS Open Access API aims, to increase the accessibility of data compiled in the AD genetics knowledge bases and encourage data reuse. This service conforms to the OpenAPI specification^47^ to harmonize information across the Open Access resources and enable programmatic access and integration of these data and annotations into analysis pipelines. Calls to the API are simple and allow users to build queries based on feature types (e.g., gene, variant, or locus), feature identifier(s) and filter criteria (e.g., experimental design or bio-source), defining an intuitive, templated interface for programmatic access. The API calls for gene or variant records return functional annotations as well as genetic evidence for AD/ADRD-risk compiled across the AD GenomicsDB, VariXam, and ADVP knowledge bases. Queries can also be made against FILER or the GenomicsDB to retrieve functional annotations and risk-associated variants within a genomic span of interest across one or multiple datasets. The API supports both single-lookups and batch queries, with paginated results to improve query and response times.

NIAGADS continues to improve our service, strengthen interoperability with other resources, and enhance the data platform to align with user feedback and new data technologies and research trends. NIAGADS plans to roll out new data discovery features for DSS using Gen3^48^. Developed by the University of Chicago, Gen3 is a mature platform that has been adopted for several other large genomics projects such as the NCI Genomic Data Commons Data Portal^49^. NIAGADS also is developing a Data Submission Portal that provides guidance and automated validation for investigators to deposit new datasets or return analysis summaries and derived data based on NIAGADS datasets. NIAGADS is working with ADSP investigators to set up a cloud-based analysis enclave. The ADSP Cloud Analysis Commons will make data on DSS directly accessible to analysts on the Amazon cloud while maintaining FISMA Moderate security, enabling investigators to collaborate more closely on the cloud and easily integrate ADSP data with other large genomics datasets.

## Acknowledgement

This work is supported by U24AG041689, U54AG052427, U01AG032984 and U01AG058654. We thank Dr. Christian J. Stoeckert, Jr., Conor Klamann, Briana Booth, NIA program officers, the NHGRI Centers for Common Disease Genomics (CCDG; formerly LSAC, Large-Scale Genome Sequencing and Analysis Centers) program and the program officer, the database of Genotypes and Phenotypes (dbGaP) team, NIH Center for Information Technology (CIT), Lumina Corps, and members of the NIA ADSP Data Access Committee (NADAC) and NIAGADS Data Use Committee for their guidance and support.

## Alzheimer’s Disease Sequencing Project (sa000001) data

The Alzheimer’s Disease Sequencing Project (ADSP) is comprised of two Alzheimer’s Disease (AD) genetics consortia and three National Human Genome Research Institute (NHGRI) funded Large Scale Sequencing and Analysis Centers (LSAC). The two AD genetics consortia are the Alzheimer’s Disease Genetics Consortium (ADGC) funded by NIA (U01 AG032984), and the Cohorts for Heart and Aging Research in Genomic Epidemiology (CHARGE) funded by NIA (R01 AG033193), the National Heart, Lung, and Blood Institute (NHLBI), other National Institute of Health (NIH) institutes and other foreign governmental and non-governmental organizations. The Discovery Phase analysis of sequence data is supported through UF1AG047133 (to Drs. Schellenberg, Farrer, Pericak-Vance, Mayeux, and Haines); U01AG049505 to Dr. Seshadri; U01AG049506 to Dr. Boerwinkle; U01AG049507 to Dr. Wijsman; and U01AG049508 to Dr. Goate and the Discovery Extension Phase analysis is supported through U01AG052411 to Dr. Goate, U01AG052410 to Dr. Pericak-Vance and U01 AG052409 to Drs. Seshadri and Fornage. Sequencing for the Follow Up Study (FUS) is supported through U01AG057659 (to Drs. PericakVance, Mayeux, and Vardarajan) and U01AG062943 (to Drs. Pericak-Vance and Mayeux). Data generation and harmonization in the Follow-up Phase is supported by U54AG052427 (to Drs. Schellenberg and Wang). The FUS Phase analysis of sequence data is supported through U01AG058589 (to Drs. Destefano, Boerwinkle, De Jager, Fornage, Seshadri, and Wijsman), U01AG058654 (to Drs. Haines, Bush, Farrer, Martin, and Pericak-Vance), U01AG058635 (to Dr. Goate), RF1AG058066 (to Drs. Haines, Pericak-Vance, and Scott), RF1AG057519 (to Drs. Farrer and Jun), R01AG048927 (to Dr. Farrer), and RF1AG054074 (to Drs. Pericak-Vance and Beecham).

The ADGC cohorts include: Adult Changes in Thought (ACT) (U01 AG006781, U19 AG066567), the Alzheimer’s Disease Research Centers (ADRC) (P30 AG062429, P30 AG066468, P30 AG062421, P30 AG066509, P30 AG066514, P30 AG066530, P30 AG066507, P30 AG066444, P30 AG066518, P30 AG066512, P30 AG066462, P30 AG072979, P30 AG072972, P30 AG072976, P30 AG072975, P30 AG072978, P30 AG072977, P30 AG066519, P30 AG062677, P30 AG079280, P30 AG062422, P30 AG066511, P30 AG072946, P30 AG062715, P30 AG072973, P30 AG066506, P30 AG066508, P30 AG066515, P30 AG072947, P30 AG072931, P30 AG066546, P20 AG068024, P20 AG068053, P20 AG068077, P20 AG068082, P30 AG072958, P30 AG072959), the Chicago Health and Aging Project (CHAP) (R01 AG11101, RC4 AG039085, K23 AG030944), Indiana Memory and Aging Study (IMAS) (R01 AG019771), Indianapolis Ibadan (R01 AG009956, P30 AG010133), the Memory and Aging Project (MAP) ( R01 AG17917), Mayo Clinic (MAYO) (R01 AG032990, U01 AG046139, R01 NS080820, RF1 AG051504, P50 AG016574), Mayo Parkinson’s Disease controls (NS039764, NS071674, 5RC2HG005605), University of Miami (R01 AG027944, R01 AG028786, R01 AG019085, IIRG09133827, A2011048), the Multi-Institutional Research in Alzheimer’s Genetic Epidemiology Study (MIRAGE) (R01 AG09029, R01 AG025259), the National Centralized Repository for Alzheimer’s Disease and Related Dementias (NCRAD) (U24 AG021886), the National Institute on Aging Late Onset Alzheimer’s Disease Family Study (NIA- LOAD) (U24 AG056270), the Religious Orders Study (ROS) (P30 AG10161, R01 AG15819), the Texas Alzheimer’s Research and Care Consortium (TARCC) (funded by the Darrell K Royal Texas Alzheimer’s Initiative), Vanderbilt University/Case Western Reserve University (VAN/CWRU) (R01 AG019757, R01 AG021547, R01 AG027944, R01 AG028786, P01 NS026630, and Alzheimer’s Association), the Washington Heights-Inwood Columbia Aging Project (WHICAP) (RF1 AG054023), the University of Washington Families (VA Research Merit Grant, NIA: P50AG005136, R01AG041797, NINDS: R01NS069719), the Columbia University Hispanic Estudio Familiar de Influencia Genetica de Alzheimer (EFIGA) (RF1 AG015473), the University of Toronto (UT) (funded by Wellcome Trust, Medical Research Council, Canadian Institutes of Health Research), and Genetic Differences (GD) (R01 AG007584). The CHARGE cohorts are supported in part by National Heart, Lung, and Blood Institute (NHLBI) infrastructure grant HL105756 (Psaty), RC2HL102419 (Boerwinkle) and the neurology working group is supported by the National Institute on Aging (NIA) R01 grant AG033193.

The CHARGE cohorts participating in the ADSP include the following: Austrian Stroke Prevention Study (ASPS), ASPS-Family study, and the Prospective Dementia Registry-Austria (ASPS/PRODEM-Aus), the Atherosclerosis Risk in Communities (ARIC) Study, the Cardiovascular Health Study (CHS), the Erasmus Rucphen Family Study (ERF), the Framingham Heart Study (FHS), and the Rotterdam Study (RS). ASPS is funded by the Austrian Science Fond (FWF) grant number P20545-P05 and P13180 and the Medical University of Graz. The ASPS-Fam is funded by the Austrian Science Fund (FWF) project I904), the EU Joint Programme – Neurodegenerative Disease Research (JPND) in frame of the BRIDGET project (Austria, Ministry of Science) and the Medical University of Graz and the Steiermärkische Krankenanstalten Gesellschaft. PRODEM-Austria is supported by the Austrian Research Promotion agency (FFG) (Project No. 827462) and by the Austrian National Bank (Anniversary Fund, project 15435. ARIC research is carried out as a collaborative study supported by NHLBI contracts (HHSN268201100005C, HHSN268201100006C, HHSN268201100007C, HHSN268201100008C, HHSN268201100009C, HHSN268201100010C, HHSN268201100011C, and HHSN268201100012C). Neurocognitive data in ARIC is collected by U01 2U01HL096812, 2U01HL096814, 2U01HL096899, 2U01HL096902, 2U01HL096917 from the NIH (NHLBI, NINDS, NIA and NIDCD), and with previous brain MRI examinations funded by R01-HL70825 from the NHLBI. CHS research was supported by contracts HHSN268201200036C, HHSN268200800007C, N01HC55222, N01HC85079, N01HC85080, N01HC85081, N01HC85082, N01HC85083, N01HC85086, and grants U01HL080295 and U01HL130114 from the NHLBI with additional contribution from the National Institute of Neurological Disorders and Stroke (NINDS). Additional support was provided by R01AG023629, R01AG15928, and R01AG20098 from the NIA. FHS research is supported by NHLBI contracts N01-HC-25195 and HHSN268201500001I. This study was also supported by additional grants from the NIA (R01s AG054076, AG049607 and AG033040 and NINDS (R01 NS017950). The ERF study as a part of EUROSPAN (European Special Populations Research Network) was supported by European Commission FP6 STRP grant number 018947 (LSHG-CT-2006-01947) and also received funding from the European Community’s Seventh Framework Programme (FP7/2007-2013)/grant agreement HEALTH-F4- 2007-201413 by the European Commission under the programme “Quality of Life and Management of the Living Resources” of 5th Framework Programme (no. QLG2-CT-2002- 01254). High-throughput analysis of the ERF data was supported by a joint grant from the Netherlands Organization for Scientific Research and the Russian Foundation for Basic Research (NWO-RFBR 047.017.043). The Rotterdam Study is funded by Erasmus Medical Center and Erasmus University, Rotterdam, the Netherlands Organization for Health Research and Development (ZonMw), the Research Institute for Diseases in the Elderly (RIDE), the Ministry of Education, Culture and Science, the Ministry for Health, Welfare and Sports, the European Commission (DG XII), and the municipality of Rotterdam. Genetic data sets are also supported by the Netherlands Organization of Scientific Research NWO Investments (175.010.2005.011, 911-03-012), the Genetic Laboratory of the Department of Internal Medicine, Erasmus MC, the Research Institute for Diseases in the Elderly (014-93-015; RIDE2), and the Netherlands Genomics Initiative (NGI)/Netherlands Organization for Scientific Research (NWO) Netherlands Consortium for Healthy Aging (NCHA), project 050-060-810. All studies are grateful to their participants, faculty and staff. The content of these manuscripts is solely the responsibility of the authors and does not necessarily represent the official views of the National Institutes of Health or the U.S. Department of Health and Human Services.

The FUS cohorts include: the Alzheimer’s Disease Research Centers (ADRC) (P30 AG062429, P30 AG066468, P30 AG062421, P30 AG066509, P30 AG066514, P30 AG066530, P30 AG066507, P30 AG066444, P30 AG066518, P30 AG066512, P30 AG066462, P30 AG072979, P30 AG072972, P30 AG072976, P30 AG072975, P30 AG072978, P30 AG072977, P30 AG066519, P30 AG062677, P30 AG079280, P30 AG062422, P30 AG066511, P30 AG072946, P30 AG062715, P30 AG072973, P30 AG066506, P30 AG066508, P30 AG066515, P30 AG072947, P30 AG072931, P30 AG066546, P20 AG068024, P20 AG068053, P20 AG068077, P20 AG068082, P30 AG072958, P30 AG072959), Alzheimer’s Disease Neuroimaging Initiative (ADNI) (U19AG024904), Amish Protective Variant Study (RF1AG058066), Cache County Study (R01AG11380, R01AG031272, R01AG21136, RF1AG054052), Case Western Reserve University Brain Bank (CWRUBB) (P50AG008012), Case Western Reserve University Rapid Decline (CWRURD) (RF1AG058267, NU38CK000480), CubanAmerican Alzheimer’s Disease Initiative (CuAADI) (3U01AG052410), Estudio Familiar de Influencia Genetica en Alzheimer (EFIGA) (5R37AG015473, RF1AG015473, R56AG051876), Genetic and Environmental Risk Factors for Alzheimer Disease Among African Americans Study (GenerAAtions) (2R01AG09029, R01AG025259, 2R01AG048927), Gwangju Alzheimer and Related Dementias Study (GARD) (U01AG062602), Hillblom Aging Network (2014-A-004-NET, R01AG032289, R01AG048234), Hussman Institute for Human Genomics Brain Bank (HIHGBB) (R01AG027944, Alzheimer’s Association “Identification of Rare Variants in Alzheimer Disease”), Ibadan Study of Aging (IBADAN) (5R01AG009956), Longevity Genes Project (LGP) and LonGenity (R01AG042188, R01AG044829, R01AG046949, R01AG057909, R01AG061155, P30AG038072), Mexican Health and Aging Study (MHAS) (R01AG018016), Multi-Institutional Research in Alzheimer’s Genetic Epidemiology (MIRAGE) (2R01AG09029, R01AG025259, 2R01AG048927), Northern Manhattan Study (NOMAS) (R01NS29993), Peru Alzheimer’s Disease Initiative (PeADI) (RF1AG054074), Puerto Rican 1066 (PR1066) (Wellcome Trust (GR066133/GR080002), European Research Council (340755)), Puerto Rican Alzheimer Disease Initiative (PRADI) (RF1AG054074), Reasons for Geographic and Racial Differences in Stroke (REGARDS) (U01NS041588), Research in African American Alzheimer Disease Initiative (REAAADI) (U01AG052410), the Religious Orders Study (ROS) (P30 AG10161, P30 AG72975, R01 AG15819, R01 AG42210), the RUSH Memory and Aging Project (MAP) (R01 AG017917, R01 AG42210Stanford Extreme Phenotypes in AD (R01AG060747), University of Miami Brain Endowment Bank (MBB), University of Miami/Case Western/North Carolina A&T African American (UM/CASE/NCAT) (U01AG052410, R01AG028786), and Wisconsin Registry for Alzheimer’s Prevention (WRAP) (R01AG027161 and R01AG054047).

The four LSACs are: the Human Genome Sequencing Center at the Baylor College of Medicine (U54 HG003273), the Broad Institute Genome Center (U54HG003067), The American Genome Center at the Uniformed Services University of the Health Sciences (U01AG057659), and the Washington University Genome Institute (U54HG003079). Genotyping and sequencing for the ADSP FUS is also conducted at John P. Hussman Institute for Human Genomics (HIHG) Center for Genome Technology (CGT).

Biological samples and associated phenotypic data used in primary data analyses were stored at Study Investigators institutions, and at the National Centralized Repository for Alzheimer’s Disease and Related Dementias (NCRAD, U24AG021886) at Indiana University funded by NIA. Associated Phenotypic Data used in primary and secondary data analyses were provided by Study Investigators, the NIA funded Alzheimer’s Disease Centers (ADCs), and the National Alzheimer’s Coordinating Center (NACC, U24AG072122) and the National Institute on Aging Genetics of Alzheimer’s Disease Data Storage Site (NIAGADS, U24AG041689) at the University of Pennsylvania, funded by NIA. Harmonized phenotypes were provided by the ADSP Phenotype Harmonization Consortium (ADSP-PHC), funded by NIA (U24 AG074855, U01 AG068057 and R01 AG059716) and Ultrascale Machine Learning to Empower Discovery in Alzheimer’s Disease Biobanks (AI4AD, U01 AG068057). This research was supported in part by the Intramural Research Program of the National Institutes of health, National Library of Medicine. Contributors to the Genetic Analysis Data included Study Investigators on projects that were individually funded by NIA, and other NIH institutes, and by private U.S. organizations, or foreign governmental or nongovernmental organizations.

The ADSP Phenotype Harmonization Consortium (ADSP-PHC) is funded by NIA (U24 AG074855, U01 AG068057 and R01 AG059716). The harmonized cohorts within the ADSP-PHC include: the Anti-Amyloid Treatment in Asymptomatic Alzheimer’s study (A4 Study), a secondary prevention trial in preclinical Alzheimer’s disease, aiming to slow cognitive decline associated with brain amyloid accumulation in clinically normal older individuals. The A4 Study is funded by a public-private-philanthropic partnership, including funding from the National Institutes of Health-National Institute on Aging, Eli Lilly and Company, Alzheimer’s Association, Accelerating Medicines Partnership, GHR Foundation, an anonymous foundation and additional private donors, with in-kind support from Avid and Cogstate. The companion observational Longitudinal Evaluation of Amyloid Risk and Neurodegeneration (LEARN) Study is funded by the Alzheimer’s Association and GHR Foundation. The A4 and LEARN Studies are led by Dr. Reisa Sperling at Brigham and Women’s Hospital, Harvard Medical School and Dr. Paul Aisen at the Alzheimer’s Therapeutic Research Institute (ATRI), University of Southern California. The A4 and LEARN Studies are coordinated by ATRI at the University of Southern California, and the data are made available through the Laboratory for Neuro Imaging at the University of Southern California. The participants screening for the A4 Study provided permission to share their de-identified data in order to advance the quest to find a successful treatment for Alzheimer’s disease. We would like to acknowledge the dedication of all the participants, the site personnel, and all of the partnership team members who continue to make the A4 and LEARN Studies possible. The complete A4 Study Team list is available on: a4study.org/a4-study-team.; the Adult Changes in Thought study (ACT), U01 AG006781, U19 AG066567; Alzheimer’s Disease Neuroimaging Initiative (ADNI): Data collection and sharing for this project was funded by the Alzheimer’s Disease Neuroimaging Initiative (ADNI) (National Institutes of Health Grant U01 AG024904) and DOD ADNI (Department of Defense award number W81XWH-12-2-0012). ADNI is funded by the National Institute on Aging, the National Institute of Biomedical Imaging and Bioengineering, and through generous contributions from the following: AbbVie, Alzheimer’s Association; Alzheimer’s Drug Discovery Foundation; Araclon Biotech; BioClinica, Inc.; Biogen; Bristol-Myers Squibb Company; CereSpir, Inc.; Cogstate; Eisai Inc.; Elan Pharmaceuticals, Inc.; Eli Lilly and Company; EuroImmun; F. Hoffmann-La Roche Ltd and its affiliated company Genentech, Inc.; Fujirebio; GE Healthcare; IXICO Ltd.;Janssen Alzheimer Immunotherapy Research & Development, LLC.; Johnson & Johnson Pharmaceutical Research & Development LLC.; Lumosity; Lundbeck; Merck & Co., Inc.;Meso Scale Diagnostics, LLC.; NeuroRx Research; Neurotrack Technologies; Novartis Pharmaceuticals Corporation; Pfizer Inc.; Piramal Imaging; Servier; Takeda Pharmaceutical Company; and Transition Therapeutics. The Canadian Institutes of Health Research is providing funds to support ADNI clinical sites in Canada. Private sector contributions are facilitated by the Foundation for the National Institutes of Health (www.fnih.org). The grantee organization is the Northern California Institute for Research and Education, and the study is coordinated by the Alzheimer’s Therapeutic Research Institute at the University of Southern California. ADNI data are disseminated by the Laboratory for Neuro Imaging at the University of Southern California; Estudio Familiar de Influencia Genetica en Alzheimer (EFIGA): 5R37AG015473, RF1AG015473, R56AG051876; Memory & Aging Project at Knight Alzheimer’s Disease Research Center (MAP at Knight ADRC): The Memory and Aging Project at the Knight-ADRC (Knight-ADRC). This work was supported by the National Institutes of Health (NIH) grants R01AG064614, R01AG044546, RF1AG053303, RF1AG058501, U01AG058922 and R01AG064877 to Carlos Cruchaga. The recruitment and clinical characterization of research participants at Washington University was supported by NIH grants P30AG066444, P01AG03991, and P01AG026276. Data collection and sharing for this project was supported by NIH grants RF1AG054080, P30AG066462, R01AG064614 and U01AG052410. We thank the contributors who collected samples used in this study, as well as patients and their families, whose help and participation made this work possible. This work was supported by access to equipment made possible by the Hope Center for Neurological Disorders, the Neurogenomics and Informatics Center (NGI: https://neurogenomics.wustl.edu/) and the Departments of Neurology and Psychiatry at Washington University School of Medicine; National Alzheimer’s Coordinating Center (NACC): The NACC database is funded by NIA/NIH Grant U24 AG072122. NACC data are contributed by the NIA-funded ADRCs: P30 AG062429 (PI James Brewer, MD, PhD), P30 AG066468 (PI Oscar Lopez, MD), P30 AG062421 (PI Bradley Hyman, MD, PhD), P30 AG066509 (PI Thomas Grabowski, MD), P30 AG066514 (PI Mary Sano, PhD), P30 AG066530 (PI Helena Chui, MD), P30 AG066507 (PI Marilyn Albert, PhD), P30 AG066444 (PI John Morris, MD), P30 AG066518 (PI Jeffrey Kaye, MD), P30 AG066512 (PI Thomas Wisniewski, MD), P30 AG066462 (PI Scott Small, MD), P30 AG072979 (PI David Wolk, MD), P30 AG072972 (PI Charles DeCarli, MD), P30 AG072976 (PI Andrew Saykin, PsyD), P30 AG072975 (PI David Bennett, MD), P30 AG072978 (PI Neil Kowall, MD), P30 AG072977 (PI Robert Vassar, PhD), P30 AG066519 (PI Frank LaFerla, PhD), P30 AG062677 (PI Ronald Petersen, MD, PhD), P30 AG079280 (PI Eric Reiman, MD), P30 AG062422 (PI Gil Rabinovici, MD), P30 AG066511 (PI Allan Levey, MD, PhD), P30 AG072946 (PI Linda Van Eldik, PhD), P30 AG062715 (PI Sanjay Asthana, MD, FRCP), P30 AG072973 (PI Russell Swerdlow, MD), P30 AG066506 (PI Todd Golde, MD, PhD), P30 AG066508 (PI Stephen Strittmatter, MD, PhD), P30 AG066515 (PI Victor Henderson, MD, MS), P30 AG072947 (PI Suzanne Craft, PhD), P30 AG072931 (PI Henry Paulson, MD, PhD), P30 AG066546 (PI Sudha Seshadri, MD), P20 AG068024 (PI Erik Roberson, MD, PhD), P20 AG068053 (PI Justin Miller, PhD), P20 AG068077 (PI Gary Rosenberg, MD), P20 AG068082 (PI Angela Jefferson, PhD), P30 AG072958 (PI Heather Whitson, MD), P30 AG072959 (PI James Leverenz, MD); National Institute on Aging Alzheimer’s Disease Family Based Study (NIA-AD FBS): U24 AG056270; Religious Orders Study (ROS): P30AG10161,R01AG15819, R01AG42210; Memory and Aging Project (MAP - Rush): R01AG017917, R01AG42210; Minority Aging Research Study (MARS): R01AG22018, R01AG42210; Washington Heights/Inwood Columbia Aging Project (WHICAP): RF1 AG054023;and Wisconsin Registry for Alzheimer’s Prevention (WRAP): R01AG027161 and R01AG054047. Additional acknowledgments include the National Institute on Aging Genetics of Alzheimer’s Disease Data Storage Site (NIAGADS, U24AG041689) at the University of Pennsylvania, funded by NIA.

## Alzheimer’s Disease Neuroimaging Initiative (sa000002) data

Data collection and sharing for this project was funded by the Alzheimer’s Disease Neuroimaging Initiative (ADNI) (National Institutes of Health Grant U01 AG024904) and DOD ADNI (Department of Defense award number W81XWH-12-2-0012). ADNI is funded by the National Institute on Aging, the National Institute of Biomedical Imaging and Bioengineering, and through generous contributions from the following: AbbVie, Alzheimer’s Association; Alzheimer’s Drug Discovery Foundation; Araclon Biotech; BioClinica, Inc.; Biogen; Bristol-Myers Squibb Company; CereSpir, Inc.; Cogstate; Eisai Inc.; Elan Pharmaceuticals, Inc.; Eli Lilly and Company; EuroImmun; F. Hoffmann-La Roche Ltd and its affiliated company Genentech, Inc.; Fujirebio; GE Healthcare; IXICO Ltd.; Janssen Alzheimer Immunotherapy Research & Development, LLC.; Johnson & Johnson Pharmaceutical Research & Development LLC.; Lumosity; Lundbeck; Merck & Co., Inc.; Meso Scale Diagnostics, LLC.; NeuroRx Research; Neurotrack Technologies; Novartis Pharmaceuticals Corporation; Pfizer Inc.; Piramal Imaging; Servier; Takeda Pharmaceutical Company; and Transition Therapeutics. The Canadian Institutes of Health Research is providing funds to support ADNI clinical sites in Canada. Private sector contributions are facilitated by the Foundation for the National Institutes of Health (www.fnih.org). The grantee organization is the Northern California Institute for Research and Education, and the study is coordinated by the Alzheimer’s Therapeutic Research Institute at the University of Southern California. ADNI data are disseminated by the Laboratory for Neuro Imaging at the University of Southern California.

### Alzheimer’s Disease Genetics Consortium (sa000003) data: ADGC_AA_WES (snd10003) data

NIH grants supported enrollment and data collection for the individual studies including: GenerAAtions R01AG20688 (PI M. Daniele Fallin, PhD); Miami/Duke R01 AG027944, R01 AG028786 (PI Margaret A. Pericak-Vance, PhD); NC A&T P20 MD000546, R01 AG28786-01A1

(PI Goldie S. Byrd, PhD); Case Western (PI Jonathan L. Haines, PhD); MIRAGE R01 AG009029 (PI Lindsay A. Farrer, PhD); ROS P30AG10161, R01AG15819, R01AG30146, TGen (PI David A. Bennett, MD); MAP R01AG17917, R01AG15819, TGen (PI David A. Bennett, MD); MARS R01AG022018 (PI Lisa L. Barnes).<u>[CL1] [KA2]</u> The NACC database is funded by NIA/NIH Grant U24 AG072122. NACC data are contributed by the NIA-funded ADCs: P30 AG019610 (PI Eric Reiman, MD), P30 AG013846 (PI Neil Kowall, MD), P30 AG062428-01 (PI James Leverenz, MD) P50 AG008702 (PI Scott Small, MD), P50 AG025688 (PI Allan Levey, MD, PhD), P50 AG047266 (PI Todd Golde, MD, PhD), P30 AG010133 (PI Andrew Saykin, PsyD), P50 AG005146 (PI Marilyn Albert, PhD), P30 AG062421-01 (PI Bradley Hyman, MD, PhD), P30 AG062422-01 (PI Ronald Petersen, MD, PhD), P50 AG005138 (PI Mary Sano, PhD), P30 AG008051 (PI Thomas Wisniewski, MD), P30 AG013854 (PI Robert Vassar, PhD), P30 AG008017 (PI Jeffrey Kaye, MD), P30 AG010161 (PI David Bennett, MD), P50 AG047366 (PI Victor Henderson, MD, MS), P30 AG010129 (PI Charles DeCarli, MD), P50 AG016573 (PI Frank LaFerla, PhD), P30 AG062429-01(PI James Brewer, MD, PhD), P50 AG023501 (PI Bruce Miller, MD), P30 AG035982 (PI Russell Swerdlow, MD), P30 AG028383 (PI Linda Van Eldik, PhD), P30 AG053760 (PI Henry Paulson, MD, PhD), P30 AG010124 (PI John Trojanowski, MD, PhD), P50 AG005133 (PI Oscar Lopez, MD), P50 AG005142 (PI Helena Chui, MD), P30 AG012300 (PI Roger Rosenberg, MD), P30 AG049638 (PI Suzanne Craft, PhD), P50 AG005136 (PI Thomas Grabowski, MD), P30 AG062715-01 (PI Sanjay Asthana, MD, FRCP), P50 AG005681 (PI John Morris, MD), P50 AG047270 (PI Stephen Strittmatter, MD, PhD).

**ADGC-TARCC-WGS** (snd10030) data: This study was made possible by the Texas Alzheimer’s Research and Care Consortium (TARCC) funded by the state of Texas through the Texas Council on Alzheimer’s Disease and Related Disorders and the Darrell K Royal Texas Alzheimer’s Initiative.

## The Familial Alzheimer Sequencing Project (sa000004) data

This work was supported by grants from the National Institutes of Health (R01AG044546, P01AG003991, RF1AG053303, R01AG058501, U01AG058922, RF1AG058501 and R01AG057777). The recruitment and clinical characterization of research participants at Washington University were supported by NIH P50 AG05681, P01 AG03991, and P01 AG026276. This work was supported by access to equipment made possible by the Hope Center for Neurological Disorders, and the Departments of Neurology and Psychiatry at Washington University School of Medicine.

We thank the contributors who collected samples used in this study, as well as patients and their families, whose help and participation made this work possible. This work was supported by access to equipment made possible by the Hope Center for Neurological Disorders, and the Departments of Neurology and Psychiatry at Washington University School of Medicine

## Brkanac- Family-based genome scan for AAO of LOAD (sa000005) data

This work was partially supported by grant funding from NIH R01 AG039700 and NIH P50 AG005136. Subjects and samples used here were originally collected with grant funding from NIH U24 AG026395, U24 AG021886, P50 AG008702, P01 AG007232, R37 AG015473, P30 AG028377, P50 AG05128, P50 AG16574, P30 AG010133, P50 AG005681, P01 AG003991, U01MH046281, U01 MH046290 and U01 MH046373. The funders had no role in study design, analysis or preparation of the manuscript. The authors declare no competing interests.

## HIHG Miami Families with AD (sa000006) data

This work was supported by the National Institutes of Health (R01 AG027944, R01 AG028786 to MAPV, R01 AG019085 to JLH, P20 MD000546); a joint grant from the Alzheimer’s Association (SG-14-312644) and the Fidelity Biosciences Research Initiative to MAPV; the BrightFocus Foundation (A2011048 to MAPV). NIA-LOAD Family-Based Study supported the collection of samples used in this study through NIH grants U24 AG026395 and R01 AG041797 and the MIRAGE cohort was supported through the NIH grants R01 AG025259 and R01 AG048927. We thank contributors, including the Alzheimer’s disease Centers who collected samples used in this study, as well as patients and their families, whose help and participation made this work possible. Study design: HNC, BWK, JLH, MAPV; Sample collection: MLC, JMV, RMC, LAF, JLH, MAPV; Whole exome sequencing and Sanger sequencing: SR, PLW; Sequencing data analysis: HNC, BWK, KLHN, SR, MAK, JRG, ERM, GWB, MAPV; Statistical analysis: BWK, KLHN, JMJ, MAPV; Preparation of manuscript: HNC, BWK. The authors jointly discussed the experimental results throughout the duration of the study. All authors read and approved the final manuscript.

## Washington Heights/Inwood Columbia Aging Project (sa000007) data

Data collection and sharing for this project was supported by the Washington Heights-Inwood Columbia Aging Project (WHICAP, PO1AG07232, R01AG037212, RF1AG054023) funded by the National Institute on Aging (NIA) and by the National Center for Advancing Translational Sciences, National Institutes of Health, through Grant Number UL1TR001873. This manuscript has been reviewed by WHICAP investigators for scientific content and consistency of data interpretation with previous WHICAP Study publications. We acknowledge the WHICAP study participants and the WHICAP research and support staff for their contributions to this study.

## Corticobasal Degeneration Study (sa000009) data

CBD Solutions funded the WES, data processing, and analysis. Assembled samples are from University College London (John Hardy), Mayo Clinic Jacksonville (Dennis Dickson), University of Pennsylvania (John Trojanowski), Emory University (Marla Gearing), Johns Hopkins University (Alex Pantelyat), Indiana University (Bernadino Ghetti), New York Brain Bank (Jean Paul Vonsattel), McClean Brain Bank (Elaine Benes), University of Texas Southwestern (Charles White), University of California Los Angeles (William Tourtelloute), and European collaborators at University Munich and Neurobiobank Munich (Gunter Hoglinger, Ulrich Muller, Hans Kretzschmr), Newcastle University, University of Barcelona (Charles Gaig), MRC London Brain Bank, Australian Brain Bank, and the University of Madrid (Alberto Rábano Gutiérrez).

## Progressive Supranuclear Palsy Study (sa000010) data

This work was funded by the following NIH grants: P01 AG017586 (VM-YL, GDS, JQT), U54 NS100693 (OR, DD, GDS), UG3 NS104095 (GDS, L-SW, OR), U54 AG052427 (l-SW, GDS), P30 AG010133 (B.G.), R01 AG057516 (AC, AM, AW, JAP, SG), R01 HL143790 (AC, SG), R01 HG010067 (SG), RF1 AG055477 (CB), P01 AG017586 (VM-YL, GDS, JQT, VMV), UG3 NS104095 and CWOW grant U54 NS100693 (DD), AG025688 and NS055077 (MG), P30 AG012300 (CLW), P30 AG053760 (APL and RA), 1P50NS091856 (RA), 5 P50 AG005134 (MPF), AG005131 (DRG), Johns Hopkins University Morris K. Udall Parkinson’s Disease Research Center of Excellence grant P50 NS038377 and Alzheimer’s Disease Research Center grant P50 AG05146 (JCT), U24 NS072026 and P30 AG19610 (TGB). This work was also funded by Cure PSP (GDS), the Rainwater Foundation (GDS), the Daniel B. Burke Endowed Chair for Diabetes Research (SG), the CHOP Center for Spatial and Functional Genomics (AW, SFG), a CUREPSP research grant (Cure PSP Grant # 515-14; 2013-2015) to P.P., the Reta Lila Weston Trust for Medical ResearFAch, the PSP Association (RdS), and the Michael J. Fox Foundation for Parkinson’s Research (TGB). G. Höglinger was funded by the German Research Foundation (DFG) under Germany’s Excellence Strategy within the framework of the Munich Cluster for Systems Neurology (EXC 2145 SyNergy – ID 390857198), the German Federal Ministry of Education and Research (BMBF, 01KU1403A EpiPD; 01EK1605A HitTau), and the NOMIS foundation (FTLD project). J. Hardy was partly funded by UKDRI limited which receives its funding from the MRC, the Alzheimer’s Society and Alzheimer Research UK. The London Neurodegenerative Diseases Brain Bank receives funding from the UK Medical Research Council (MR/L016397/1) and as part of the Brains for Dementia Research programme, jointly funded by Alzheimer’s Research UK and the Alzheimer’s Society. Queen Square Brain Bank is supported by the Reta Lila Weston Institute for Neurological Studies and the Medical Research Council UK. Newcastle Brain Tissue Resource is funded in part by a grant from the UK Medical Research Council (MR/L016451/1) and by Brains for Dementia Research, a joint venture between Alzheimer’s Society and Alzheimer’s Research UK (CMM) and National Institute of Health Research Biomedical Research Centre at Newcastle upon Tyne Hospitals NHS Foundation Trust and Newcastle University (CMM). This work was partly funded by UKDRI limited which receives its funding from the MRC, the Alzheimer’s Society and Alzheimer Research UK (JH). The Mayo Clinic Florida had support from a Morris K. Udall Parkinson’s Disease Research Center of Excellence (NINDS P50 #NS072187), CurePSP and the Tau Consortium. OAR is supported by a NINDS Tau Center without Walls (U54-NS100693), NINDS R01-NS078086 and the Mayo Clinic Center for Individualized Medicine. Funding provided by CurePSP through the generous support of the Peebler PSP Research Foundation in memory of Charles D. Peebler Jr. and Drs. Jeffrey S. and Jennifer R. Friedman in memory of Morton L. Friedman.

## Accelerating Medicines Partnership-Alzheimer’s Disease (AMP-AD) (sa000011) data

**Mayo RNAseq Study-** Study data were provided by the following sources: The Mayo Clinic Alzheimer’s Disease Genetic Studies, led by Dr. Nilufer Ertekin-Taner and Dr. Steven G. Younkin, Mayo Clinic, Jacksonville, FL using samples from the Mayo Clinic Study of Aging, the Mayo Clinic Alzheimer’s Disease Research Center, and the Mayo Clinic Brain Bank. Data collection was supported through funding by NIA grants P50 AG016574, R01 AG032990, U01 AG046139, R01 AG018023, U01 AG006576, U01 AG006786, R01 AG025711, R01 AG017216, R01 AG003949, NINDS grant R01 NS080820, CurePSP Foundation, and support from Mayo Foundation. Study data includes samples collected through the Sun Health Research Institute Brain and Body Donation Program of Sun City, Arizona. The Brain and Body Donation Program is supported by the National Institute of Neurological Disorders and Stroke (U24 NS072026 National Brain and Tissue Resource for Parkinson’s Disease and Related Disorders), the National Institute on Aging (P30 AG19610 Arizona Alzheimer’s Disease Core Center), the Arizona Department of Health Services (contract 211002, Arizona Alzheimer’s Research Center), the Arizona Biomedical Research Commission (contracts 4001, 0011, 05-901 and 1001 to the Arizona Parkinson’s Disease Consortium) and the Michael J. Fox Foundation for Parkinson’s Research

**ROSMAP**- We are grateful to the participants in the Religious Order Study, the Memory and Aging Project. This work is supported by the US National Institutes of Health [U01 AG046152, R01 AG043617, R01 AG042210, R01 AG036042, R01 AG036836, R01 AG032990, R01 AG18023, RC2 AG036547, P50 AG016574, U01 ES017155, KL2 RR024151, K25 AG041906- 01, R01 AG30146, P30 AG10161, R01 AG17917, R01 AG15819, K08 AG034290, P30 AG10161 and R01 AG11101.

**Mount Sinai Brain Bank (MSBB)**- This work was supported by the grants R01AG046170, RF1AG054014, RF1AG057440 and R01AG057907 from the NIH/National Institute on Aging (NIA). R01AG046170 is a component of the AMP-AD Target Discovery and Preclinical Validation Project. Brain tissue collection and characterization was supported by NIH HHSN271201300031C.

## University of Pittsburgh- Kamboh (sa000012) data

This study was supported by the National Institute on Aging (NIA) grants AG030653, AG041718, AG064877 and P30-AG066468.

## NACC Genentech Study (sa000013) data

We would like to thank study participants, their families, and the sample collectors for their invaluable contributions. This research was supported in part by the National Institute on Aging grant U01AG049508 (PI Alison M. Goate). This research was supported in part by Genentech, Inc. (PI Alison M. Goate, Robert R. Graham).

The NACC database is funded by NIA/NIH Grant U01 AG016976. NACC data are contributed by these NIA-funded ADCs: P30 AG013846 (PI Neil Kowall, MD), P50 AG008702 (PI Scott Small, MD), P50 AG025688 (PI Allan Levey, MD, PhD), P30 AG010133 (PI Andrew Saykin, PsyD), P50 AG005146 (PI Marilyn Albert, PhD), P50 AG005134 (PI Bradley Hyman, MD, PhD), P50 AG016574 (PI Ronald Petersen, MD, PhD), P30 AG013854 (PI M. Marsel Mesulam, MD), P30 AG008017 (PI Jeffrey Kaye, MD), P30 AG010161 (PI David Bennett, MD), P30 AG010129 (PI Charles DeCarli, MD), P50 AG016573 (PI Frank LaFerla, PhD), P50 AG005131 (PI Douglas Galasko, MD), P30 AG028383 (PI Linda Van Eldik, PhD), P30 AG010124 (PI John Trojanowski, MD, PhD), P50 AG005142 (PI Helena Chui, MD), P30 AG012300 (PI Roger Rosenberg, MD), P50 AG005136 (PI Thomas Grabowski, MD), P50 AG005681 (PI John Morris, MD), P30 AG028377 (Kathleen Welsh-Bohmer, PhD), and P50 AG008671 (PI Henry Paulson, MD, PhD).

Samples from the National Cell Repository for Alzheimer’s Disease (NCRAD), which receives government support under a cooperative agreement grant (U24 AG21886) awarded by the National Institute on Aging (NIA), were used in this study. We thank contributors who collected samples used in this study, as well as patients and their families, whose help and participation made this work possible.

The Alzheimer’s Disease Genetics Consortium supported the collection of samples used in this study through National Institute on Aging (NIA) grants U01AG032984 and RC2AG036528.

## Cache County Study (sa000014) data

We acknowledge the generous contributions of the Cache County Memory Study participants. Sequencing for this study was funded by RF1AG054052 (PI: John S.K. Kauwe).

## NIH, CurePSP and Tau Consortium PSP WGS (sa000015) data

This project was funded by the NIH grant UG3NS104095 and supported by grants U54NS100693 and U54AG052427. Queen Square Brain Bank is supported by the Reta Lila Weston Institute for Neurological Studies and the Medical Research Council UK. The Mayo Clinic Florida had support from a Morris K. Udall Parkinson’s Disease Research Center of Excellence (NINDS P50 #NS072187), CurePSP and the Tau Consortium. The samples from the University of Pennsylvania are supported by NIA grant P01AG017586.

## CurePSP and Tau Consortium PSP WGS (sa000016) data

This project was funded by the Tau Consortium, Rainwater Charitable Foundation, and CurePSP. It was also supported by NINDS grant U54NS100693 and NIA grants U54NS100693 and U54AG052427. Queen Square Brain Bank is supported by the Reta Lila Weston Institute for Neurological Studies and the Medical Research Council UK. The Mayo Clinic Florida had support from a Morris K. Udall Parkinson’s Disease Research Center of Excellence (NINDS P50 #NS072187), CurePSP and the Tau Consortium. The samples from the University of Pennsylvania are supported by NIA grant P01AG017586. Tissues were received from the Victorian Brain Bank, supported by The Florey Institute of Neuroscience and Mental Health, The Alfred and the Victorian Forensic Institute of Medicine and funded in part by Parkinson’s Victoria and MND Victoria. We are grateful to the Sun Health Research Institute Brain and Body Donation Program of Sun City, Arizona for the provision of human biological materials (or specific description, e.g. brain tissue, cerebrospinal fluid). The Brain and Body Donation Program is supported by the National Institute of Neurological Disorders and Stroke (U24 NS072026 National Brain and Tissue Resource for Parkinson’s Disease and Related Disorders), the National Institute on Aging (P30 AG19610 Arizona Alzheimer’s Disease Core Center), the Arizona Department of Health Services ( contract 211002, Arizona Alzheimer’s Research Center), the Arizona Biomedical Research Commission (contracts 4001, 0011, 05- 901 and 1001 to the Arizona Parkinson’s Disease Consortium) and the Michael J. Fox Foundation for Parkinson’s Research. Biomaterial was provided by the Study Group DESCRIBE of the Clinical Research of the German Center for Neurodegenerative Diseases (DZNE).

## The Diagnostic Assessment of Dementia for the Longitudinal Aging Study of India (LASI-DAD) (sa000019) data

The Longitudinal Aging Study in India, Diagnostic Assessment of Dementia Study. Produced and distributed by the University of Southern California with funding from the National Institute on Aging (grant numbers R01AG051125 and U01AG065958), Los Angeles, CA.

## Dissecting the Genomic Etiology of non-Mendelian Early-Onset Alzheimer Disease (EOAD) and Related Phenotypes (sa000023) data

This work was supported by the National Institutes of Health (NIH) grant R01AG064614. The ADSP-FUS is supported by U01AG057659.

The National Institutes of Health, National Institute on Aging (NIH-NIA) supported this work through the following grants: ADGC, U01 AG032984, RC2 AG036528; samples from the National Centralized Repository for Alzheimer’s Disease and Related Dementias (NCRAD), which receives government support under a cooperative agreement grant (U24 AG21886) awarded by the National Institute on Aging (NIA), were used in this study. Sequencing data generation and harmonization is supported by the Genome Center for Alzheimer’s Disease, U54AG052427, and data sharing is supported by NIAGADS, U24AG041689. We thank contributors who collected samples used in this study, as well as patients and their families, whose help and participation made this work possible.

NIH grants supported enrollment and data collection for the individual studies including the Alzheimer’s Disease Centers (ADC, P30 AG062429 (PI James Brewer, MD, PhD), P30 AG066468 (PI Oscar Lopez, MD), P30 AG062421 (PI Bradley Hyman, MD, PhD), P30 AG066509 (PI Thomas Grabowski, MD), P30 AG066514 (PI Mary Sano, PhD), P30 AG066530 (PI Helena Chui, MD), P30 AG066507 (PI Marilyn Albert, PhD), P30 AG066444 (PI John Morris, MD), P30 AG066518 (PI Jeffrey Kaye, MD), P30 AG066512 (PI Thomas Wisniewski, MD), P30 AG066462 (PI Scott Small, MD), P30 AG072979 (PI David Wolk, MD), P30 AG072972 (PI Charles DeCarli, MD), P30 AG072976 (PI Andrew Saykin, PsyD), P30 AG072975 (PI David Bennett, MD), P30 AG072978 (PI Neil Kowall, MD), P30 AG072977 (PI Robert Vassar, PhD), P30 AG066519 (PI Frank LaFerla, PhD), P30 AG062677 (PI Ronald Petersen, MD, PhD), P30 AG079280 (PI Eric Reiman, MD), P30 AG062422 (PI Gil Rabinovici, MD), P30 AG066511 (PI Allan Levey, MD, PhD), P30 AG072946 (PI Linda Van Eldik, PhD), P30 AG062715 (PI Sanjay Asthana, MD, FRCP), P30 AG072973 (PI Russell Swerdlow, MD), P30 AG066506 (PI Todd Golde, MD, PhD), P30 AG066508 (PI Stephen Strittmatter, MD, PhD), P30 AG066515 (PI Victor Henderson, MD, MS), P30 AG072947 (PI Suzanne Craft, PhD), P30 AG072931 (PI Henry Paulson, MD, PhD), P30 AG066546 (PI Sudha Seshadri, MD), P20 AG068024 (PI Erik Roberson, MD, PhD), P20 AG068053 (PI Justin Miller, PhD), P20 AG068077 (PI Gary Rosenberg, MD), P20 AG068082 (PI Angela Jefferson, PhD), P30 AG072958 (PI Heather Whitson, MD), P30 AG072959 (PI James Leverenz, MD). The Miami ascertainment and research were supported in part through: RF1AG054080, R01AG027944, R01AG019085, R01AG028786-02, RC2AG036528. The Columbia ascertainment and research were supported in part through: R37AG015473 and U24AG056270. The University of Washington ascertainment and research were supported in part through R01AG044546, RF1AG053303, RF1AG058501, U01AG058922 and R01AG064877.

## Data Availability

Open-access data can be freely downloaded from the NIAGADS website. Restricted-access data are available through the NIAGADS Data Sharing Service (<u>dss.niagads.org</u>) and require an approved data access request to ensure compliance with privacy and ethical guidelines and in accordance with the terms outlined by the submitting Institutional Review Boards (IRBs) and the consent provided by research participants. For detailed instructions on accessing specific datasets, including open and restricted data, please visit the NIAGADS website at www.niagads.org.

## Alzheimer’s Disease Sequencing Project Consortium Authors

Larry D. Adams^1^, Shahzhad Ahmad^2^, Najaf Amin^3^, Shea Andrews^4^, Dagmar Bacikova^5^, Sandra Barral Rodriguez^6^, Jackie Bartlett^7^, Gary Beecham^1^, Jennifer Below^8^, Penelope Benchek^7^, Joshua Bis^9^, Elizabeth Blue^9^, Eric Boerwinkle^10^, Jose Bras^11^, Min Soo Byun^12^, Alessandra F. A. Chesi^13^, Yi-Fan Chou^14^, Jaeyoon Chung^15^, Hata Comic^2^, Ryan Corces^16^, Hannah Craft^17^, Marissa Cranney^14^, Carlos Cruchaga^18^, Mike Cuccaro^1^, Clifton Dalgard^5^, Anjali Das^19^, Christos Davatzikos^14^, Tyler Day^9^, Phil De Jager^6^, Anita DeStefano^15^, Beth Dombrosk^14^, Logan Dumitrescu^8^, Susan Dutcher^18^, Kelley Faber^17^, John Farrell^15^, Lindsay Farrer^15^, Victoria Fernandez^18^, Bernard Fongang^20^, Myriam Fornage^21^, Tatiana Foroud^17^, Robert Babak Frayabi^14^, Yun Freudenberg-Hua^22^, Brian Fulton-Howard^23^, Richard Gibbs^10^, John R. Gilbert^1^, Jungsoo Gim^24^, Alison Goate^23^, Struan F. A. Grant^13^, Michael Greicius^25^, Tony Griswold^1^, Vilmundur Guðnason^26^, Rita Guerreiro^11^, Jonathan Haines^7^, Kara Hamilton-Nelson^1^, Xudong Han^15^, Nancy Heard-Costa^15^, Timothy Hohman^8^, Andrea Horimoto^9^, Heng Huang^27^, Jack Humphrey^23^, Xueqiu Jian^20^, Taeho Jo^17^, Gyungah Jun^15^, Moon-Il Kang^15^, Sharon Kardia^28^, Keoni Kauwe^29^, David Knowles^19^, Anshul Kundaje^25^, Brian Kunkle^1^, Zainab Kurshid^15^, Nicholas Kushch^1^, Kaci Lacy^17^, Chirag Lakhani^19^, Suzanne Leal^6^, Dong Young Lee^12^, Alan J Lerner^7^, Donghe Li^15^, Teresa Lin^19^, Eugine Lin^9^, Kathryn Luetta^15^, Audrey Lynn^7^, Yiyi Ma^6^, John Malamon^14^, Edoardo Dado^23^, Logue Mark^15^, Eden Martin^1^, Richard Mayeux^6^, John McNeil^3^, Pedro Mena^1^, Jesse Mez^15^, Stephen Montgomery^25^, Tom Montine^25^, Rafael Nafikov^9^, Achal Neupane^18^, Kwangsik Nho^17^, Kelly Nudelman^17^, Karen Nuytemans^1^, Tulsi Patel^23^, Gina Peloso^15^, Peggy Pericak-Vance^1^, Achilleas Pitsillides^15^, Bruce Psaty^9^, Towfique Raj^23^, Farid Rajabli^1^, Christiane Reitz^6^, Alan Renton^23^, Dolly Reyes-Dumeyer^6^, Shannon Risacher^17^, Muralidharan Sargurupremraj^20^, Chloe Sarnowski^21^, Claudia Satizabal^20^, Andrew J Saykin^17^, Mike Schmidt^1^, Sudha Seshadri^20^, Andrew Sharp^23^, Li Shen^14^, Richard Sherva^15^, Susan Slifer^1^, Jennifer Smith^28^, Yeunjoo Song^7^, Peter St. George-Hyslop^30^, Xinyu Sun^15^, Marlene Tejeda^15^, Seth Temple^9^, Sophia Thomopolous^31^, Paul Thompson^31^, Timothy Thornton^9^, Tong Tong^15^, Cornelia van Duijn^4^, Jeffery Vance^1^, Badri Vardarajan^6^, Ricardo Vialle^23^, Sungho Wang^12^, Nicolas Wheeler^7^, Patrice Whitehead^1^, Ellen Wijsman^9^, Scott Williams^7^, Benjamin Wolozin^15^, Kim Worley^10^, Dahyun Yi^12^, Habil Zare^20^, Xiaoling Zhang^1^, Wei Zhao^28^, Congcong Zhu^15^

1. University of Miami

2. Erasmus Medical University

3. Monash University

4. University of Oxford

5. United States Uniformed Health Services

6. Columbia University

7. Case Western Reserve University

8. Vanderbilt University

9. University of Washington

10. Baylor College of Medicine

11. Van Andel Institute

12. Seoul National University

13. Children’s Hospital of Pennsylvania

14. University of Pennsylvania

15. Boston University

16. University of California, San Francisco

17. Indiana University

18. Washington University St. Louis

19. New York Genome Center

20. University of Texas San Antonio

21. University of Texas Houston

22. Northwell Health

23. Mt. Sinai School of Medicine

24. Chosun University

25. Stanford University

26. University of Iceland

27. University of Pittsburgh

28. University of Michigan

29. Brigham Young University

30. University of Toronto

31. University of Southern California

