## Supplementary material for "NIAGADS: A Comprehensive National Data Repository for Alzheimer’s Disease and Related Dementia Genetics and Genomics Research": Online Methods

**Receiving data.** All data submissions are accompanied with proper documentation including the National Institute on Aging (NIA) Genome Sharing Plan, the NIH Genome Data Sharing (GDS) policy Institutional Certification form, and a data transfer agreement when applicable with the University of Pennsylvania which operates NIAGADS through a cooperative agreement with NIA. The submitter completes a Data Registration form with data description, contact information, references, supporting grants and acknowledgement information and other accompanying metadata. File transfer is conducted using secure methods, either via File Transfer Protocol (FTP) or Amazon Simple Storage Service (S3), depending on the size and sensitivity of the datasets. Upon receiving the files and forms, NIAGADS data curators review the submissions to ensure accuracy and compliance with established data formatting standards based on genomics and AD research best practices. Unique dataset accession numbers and DOIs are assigned for each dataset. All metadata about the dataset is stored in the Data Sharing Service (DSS) backend.

**Determination of research use limitations from informed consent.** NIAGADS uses the GDS Institutional Certification (IC) to determine research use limitations for each participant in each cohort. If a cohort already exists in the NIAGADS repository, the existing Institutional Certification (IC) is used. For a new cohort, NIAGADS requests the PI and the IRB of the institution administering the cohort (sometimes different from the institution submitting the data) to complete the IC. This practice ensures that participants who are deposited into NIAGADS have consistent research use limitations across different datasets.

NIAGADS then reviews the completed IC to ensure that all necessary information and approvals are in place, and assigns a research use limitation/consent level to each participant accordingly, and whether Genomic Summary Results (GSR) such as GWAS summary statistics can be shared as open access or requires controlled access per GDS policy. For datasets that involve aggregated/summary statistics, the most restricted consent level among all participants is applied to ensure that the privacy and consent of participants are respected and that the data use complies with ethical guidelines. All IC documents are stored securely in a database for future reference and audits.

**Quality control of data.** NIAGADS verifies the integrity of the files by checking the submitted manifest of files and MD5 checksums, and cross-checks sample IDs across all files to ensure that every sample is accounted for. Corrupted files are flagged and the submitter is notified to fix any data issues. NIAGADS then reviews cohort information and consent levels, and checks if any participants have already been submitted under a different cohort name to ensure data consistency.

ADSP data quality control. For data submitted to the Alzheimer’s Disease Sequencing Project (ADSP), additional validation steps are performed. First, phenotypes and pedigree structures are reviewed to ensure accuracy and consistency. NIAGADS ensures that all variables match the data dictionary and are logically consistent: for example, age of onset of the disease should be less than the age at death for clinical diagnoses. Whole exome and whole genome sequence data are processed and quality controlled by the Genome Center for Alzheimer’s Disease (GCAD) protocol. NIAGADS releases sequencing data in compressed read alignment map (CRAM) format files, genome variant call format (gVCF) files, and cumulative joint call genotype files in VCF formats that contains genotype and quality information for every genotype in every sample in all ADSP genomes released so far. Whole genomes and whole exomes are called and released separately.

**Preparing dataset for release.** Every new dataset is assigned a unique accession number for investigators and data users to reference in publications. The NIAGADS Data Management Team reviews the datasets for HIPAA compliance (e.g. ages >90 are censored), documentation completeness, and file integrity. Next, NIAGADS compiles standardized information for the dataset including database entries, characterization of subjects and samples, a file manifest with data types, and harmonized phenotypes. Next, the data files are split by consent per individual if the dataset includes multiple levels of research use limitations**.** NIAGADS then transfers files to the DSS S3 storage and generates a sample manifest with MD5 checksums and S3 file paths and file sizes. NIAGADS creates and completes DARM and DSS WordPress pages along with dataset documentation and release notes. After another round of review by the NIAGADS Data Management Team, the dataset and web page is sent to the contributing investigator for their review. Finally, all key information and web page contents are uploaded into the DARM database for data release on the DSS home page.

**Processing Data Access Requests (DARs).** NIAGADS DSS implements a detailed approval process for DARs in full compliance with the NIH GDS policy. After a DAR is received, NIAGADS staff works with an NIH administrator to verify timely the identity of the requestor, their affiliating institution, and the Signing Official (SO) using their eRA IDs. This ensures the requestor is a qualified investigator in good standing with the institution, the Signing Official can represent the institution legally, and the institution has privilege to access restricted data from NIH and is not under embargo. Once the investigator completes the DAR, the DAR is routed to the institutional SO during the “SO Review” stage to review and certify the application on behalf of the institution. In the "Admin Review" stage, the NIAGADS staff review the DAR again before sending it to the NIH Admin to review the full DAR including verification of the IT director. If the DAR is complete, the DAR moves to the "DAC Review" stage where the Data Access Committee, made up of NIH program officials, reviews the DAR. Finally, in the "Final Review" stage, NIAGADS staff review the DAR one last time. For new Data Access Requests, a fully executed Data Transfer Agreement (DTA) is required before approval, while a new DTA is not necessary for renewal applications.

**GenomicsDB.** The AD GenomicsDB (<https://www.niagads.org/genomics>) is a publicly accessible knowledgebase featuring genetic datasets and genomic annotations related to Alzheimer's disease. It employs a specialized system architecture to implement and uphold stringent standards enabling the efficient harmonization of GWAS summary statistics datasets from the NIAGADS repository and >400M variants from the ADSP joint genotype calling efforts with functional annotations. Variant annotations, including allele frequencies, predicted variant effects, and potential deleteriousness are determined using the ADSP annotation pipeline (Butkiewicz et al., 2018; Wheeler et al., 2020). Visitors to the site can interactively mine or browse the annotated datasets on a customized genome browser, or explore interactive reports compiled in the contexts of genes or variants. All genomic features are uniquely identified using standard reference identifiers (e.g., refSNP IDs, Ensembl IDs) allowing NIAGADS to link equivalent features across disparate annotation resources and provide permalinks back to GenomicsDB reports.

The site is powered by an open-source database system and web development kit (WDK (Doherty, 2023)), developed by the Eukaryotic Pathogen, Vector and Host Informatics (VEuPathDB) Bioinformatics Resource Center (Amos et al., 2022). The VEuPathDB WDK offers a query engine that connects a big data optimized PostgreSQL database system to the website front-end and interactive JavaScript visualizations through an easily extensible XML data model and Java/Jersey REST services. The WDK leverages the data model and the EDAM Ontology of bioscientific data analysis and data management (Ison et al., 2013) to automatically generate and organize searches, search results, and reports. Full details on the database design, implementation, and contents are available in Greenfest-Allen et. al 2024(Greenfest-Allen et al., 2024).

**ADVP.** The ADVP website (<https://advp.niagads.org>) offers access to systematically organized and harmonized AD-related genetic associations as reported in publications, using a standardized metadata schema. The ADVP web server runs on Amazon Web Services (AWS) cloud computing instance (m5.4xlarge) using MySQL relational database management system as a back-end and a PHP/JQuery-based web front-end. All the publication, variant and association information stored in ADVP relational database is organized into multiple tables including publications, variant, and association tables. The web front-end provides multiple data views for publications, genes, variants, and association records including interactive chromosome ideogram-based view of association data and interactive variant viewer by population and phenotype.

ADVP consists of an extensive collection of associations curated from >200 GWAS publications from Alzheimer’s Disease Genetics Consortium (ADGC) and other consortia. Genetic associations were systematically extracted, harmonized and annotated from both the genome-wide significant and suggestive loci reported in these publications. To ensure consistent representation of AD genetic findings across publications, we designed meta-data schema for systematic curation and harmonization of genetic associations at the publication, variant and association level, thus enables us to harmonize these across publications.

**FILER.** FILER web server (<https://lisanwanglab.org/FILER>) provides access to functional genomic and annotation data tracks in FILER. To provide a modular and extensible data storage architecture, FILER tracks are stored in a separate directory for each data source. Each data source directory is then further organized by experimental assay, data type, and genomic build into sub-directories. The last level of the directories corresponds to individual data collections. Each data collection thus holds tracks of the same type and data format and is individually Giggle-indexed to provide efficient access/querying based on a single genomic region or a collection of genomic regions of interest (e.g., see ‘Search’ section on the website). Meta-information table contains the standardized information for each data track including its unique ID and a reference to the data collection directory containing the track. The storage of the meta-information for the FILER genomic data tracks was implemented using MySQL database engine, allowing to query FILER contents by one or more attributes including data source, assay, cell type and others (e.g., see ‘Browse’ section on the website). All the FILER scripts were written in PHP and Javascript for the website, and AWK and Bash scripting languages for the genomic data querying and analysis backend. Highcharts (https://api.highcharts.com/highcharts) and R (v3.5.3) were used for plotting and visualization. Genomic data indexing and querying was implemented based on the Giggle genomic search engine (CITE 29309061).

FILER integrates many publicly available functional genomic and annotation resources including ENCODE(Consortium, 2012; Davis et al., 2018; Luo et al., 2020), GTEx(Consortium, 2020; Consortium et al., 2017), Roadmap Epigenomics(Roadmap Epigenomics et al., 2015), eQTL catalogue(Kerimov et al., 2023) and others (Supplementary Table SXXX ). All individual genomic datasets from these data sources are curated, uniformly processed, and standardized into consistent BED-based formats using hipFG(Cifello et al., 2023) functional genomic data harmonization and integration pipeline. Data-source-specific metadata schemas were matched across data sources to generate standardized, consistent meta-data descriptions for each of the FILER data tracks. All data in FILER is organized into >100 data collections by primary data source, experimental assay, genomic feature type, genomic build, and other data attributes. This ensures that each data collection only contains tracks with the same schema, genome assembly, and the same experimental protocol. Each such data collection is then Giggle-indexed(Layer et al., 2018) to support genomic interval-based queries (e.g. overlaps across data collections) with high efficiency. Current FILER v1.0(Kuksa et al., 2022) provides streamlined access to >70,000 harmonized data tracks across >20 diverse data sources, >1,100 cell types/tissues and >20 experimental assay and data types. Supplementary Tables SXXX provide details on data sources and data collections available in FILER. Access to the FILER data is provided using FILER web server (https://lisanwanglab.org/FILER).

**VariXam.** The VariXam database and website offers access to quality information of called variants in the ADSP whole exome and whole genome dataset. The VariXam web server runs on AWS cloud computing instance t3.medium (two CPUs and 4Gb memory) using PostgreSQL as a back-end and Apollo and GraphQL as the web front-end. All the publication, genotype, and variant information stored in the VariXam relational database is organized into multiple tables including publications, genotype, and variant tables. The web front-end provides data views for quality information for genotypes and variants curated from the ADSP data.

**NIAGADS Open Access API.** The NIAGADS Open Access API (<https://api.niagads.org>; available in beta) meets OpenAPI standards to provide an intuitive interface for programmatic access to the NIAGADS Open Access data and annotation resources (incl., ADVP, VariXam, the AD GenomicsDB, and FILER).

The API is implemented using FAST-API, a high-performance Python web framework that utilizes type hints to generate the API's structure and documentation, while also bolstering its security through type-based validation of user requests. API calls are straightforward, utilizing templated URLs that specify a data track or feature IDs (e.g., gene symbols, refSNP IDs, genomic intervals) and filtering criteria with Boolean expressions. Results are returned in the JSON structured text format defined by formal JSON schemas. The Python backend is paired with a client-side front-end, implemented in the Next.js JavaScript framework. This layer enables users to receive API responses not only as structured text, but also as interactive tables, charts, or as tracks displayed in the customized NIAGADS genome and LocusZoom browsers that draw on reference data from the AD GenomicsDB. Communication between the server-side API and the visualization tools on the client-side is mediated by a KeyDB in-memory database that temporarily stores query results, allowing for efficient server-side processing, as well as paging and short-term storage of large user queries.

To assist users in navigating the API, we provide a generative AI interface that leverages the LangChain(Chase, 2022) framework to build a large language model (LLM) for translating natural language inquiries into API calls. The application plans and optionally carry out API calls based on user inquiries to deliver results as structured data or interactive visualizations. The LLM has been trained on the NIAGADS Open Access API specification, along with example tasks and domain-specific controlled vocabularies.
